# System-Wide Investments Enhance HIV, TB and Malaria Control in Malawi and Deliver Greater Health Impact

**DOI:** 10.1101/2025.04.29.25326667

**Authors:** Tara D Mangal, Sakshi Mohan, Margherita Molaro, Joseph Collins, Tim Colbourn, Eva Janoušková, Rachel E Murray-Watson, Dominic Nkhoma, Andrew Phillips, Bingling She, Pakwanja D Twea, Simon Walker, Paul Revill, Timothy B. Hallett

## Abstract

Global health initiatives have expanded access to treatment for infectious diseases - especially HIV, tuberculosis, and malaria (HTM) - in low- and middle-income countries. However, these “vertically”-funded programs often operate within fragile health systems, where workforce shortages and supply chain failures constrain their effectiveness and sustainability ^1,2^. Meanwhile, evaluating the health impact and value-for-money of “horizontal” investments in systems, such as supply chain strengthening or boosting healthcare workforce, and their synergies with vertical programs (through “diagonal” investments combining both) - remains challenging because their benefits are mediated through improvements in many aspects of healthcare delivery and are therefore difficult to measure^3^.

Using a dynamic microsimulation model of Malawi’s healthcare system, we show that a diagonal investment approach yields a four-fold greater health impact, measured in disability-adjusted life years (DALYs) averted, than the vertical approach. This approach not only improves health outcomes for non-HTM causes of DALYs but also amplifies the effect on DALYs caused by HTM. Additionally, diagonal investments offer greater value for money and a 24.94% higher return on investment (6.67 [5.81 - 6.85] compared with 5.34 [3.44 - 6.24]), even after accounting for their additional costs. Our findings demonstrate that HSS investments generate synergistic effects, amplifying the benefits of GHIs while also strengthening broader healthcare delivery. These results support a shift toward more integrated global health financing strategies.

## Main

Over the past thirty years, global health initiatives (GHIs) have directed substantial resources into programs targeting HIV/AIDS, tuberculosis, and malaria (HTM)^1^. These efforts, spearheaded by organizations such as the Global Fund to Fight AIDS, Tuberculosis and Malaria (Global Fund), the Global Alliance for Vaccines and Immunisation (GAVI), and the President’s Emergency Plan for AIDS Relief (PEPFAR), have significantly expanded access to essential medicines, diagnostics, and health services, resulting in large improvements in population health. By 2023, new HIV infections had dropped to their lowest levels since the 1980s, with nearly 31 million people receiving lifesaving antiretroviral therapy (ART)^4^. TB treatment coverage in Global Fund-supported countries reached 70%, from 45% in 2010^5^. Malaria control efforts prevented 2.1 billion cases and 11.7 million deaths globally between 2000 and 2022^6^. Together, these initiatives have contributed to a rise in life expectancy in sub-Saharan Africa, from 56.3 years in 2010 to 61.1 years in 2023^7^.

These disease-specific (“vertical”) investments often led to fragmented service delivery, straining fragile health systems ^8,9^. Recognizing this, GHIs began incorporating health system strengthening (HSS) into their funding strategies, aiming to enhance service delivery, supply chains, and health information systems. However, the majority of HSS funding remains tethered to disease-specific interventions rather than broader, system-wide investments^1^, often leading to inefficient parallel health system structures ^10,11^.

A major contributor to this narrow investment focus is a critical gap in empirical evidence on the health impact and efficiency of “horizontal” investments, which present an alternative to the better-evidenced vertical investments. The challenge in measuring the health impact of horizontal investments arises because their benefits are mediated through improvements in many different aspects of healthcare delivery rather than direct, easily measurable and attributable health outcomes^3^. This evidence gap and its effect of funding preferences threatens the efficiency, and effectiveness of global health investments, particularly in settings with severely constrained health systems^11^.

Recent studies highlight that health system constraints, particularly human resources for health (HRH) shortages and weak supply chains, can impede the scalability of HTM programs ^12–14^. Evidence increasingly points to the synergistic value of horizontal and vertical investments ^15,16^. So-called “diagonal approaches”^17^ which integrate HRH strengthening, improved availability of consumables, and disease-specific interventions, have been shown to enhance both efficiency and equity, yielding substantially greater health benefits than isolated approaches ^18,19^. Despite this, horizontal and diagonal approaches are underexplored, particularly in high-burden LMIC settings where health system challenges are most acute.

Malawi exemplifies the challenges faced by countries with constrained health systems. Severe shortages of HRH ^20–22^, along with fragile supply chains and frequent stock-outs of essential consumables ^23,24^, undermine service delivery and limit the scalability of health interventions and impact^14^. Compounding these challenges, Malawi bears a high burden of both HTM and non-HTM diseases, such as acute lower respiratory infections (ALRI) and diarrheal diseases, which disproportionately affect marginalized populations^25^.

Here we developed a dynamic modelling framework to assess the impact of alternative investment strategies on healthcare service delivery and disease burden in Malawi, explicitly accounting for real-world constraints such as workforce shortages and supply chain disruptions. Utilizing the *Thanzi La Onse* (TLO) model^24^, an individual-based simulation of population health and healthcare interactions, we evaluated three investment approaches: vertical (HTM), horizontal (HSS), and diagonal (HTM integrated with HSS). This represents the first quantitative assessment of the health impact and value-for-money of horizontal HSS investments, their comparison with vertical investments, and the potential synergies when these approaches are combined.

To conduct our analysis, we projected the cost and health impact of a series of alternative investment approaches over an 11-year period (2025 - 2035), compared to a *Baseline* scenario where health system capacity and the coverage targets of HTM disease programs remain at 2024 levels. The vertical approach encompassed expanded HIV, TB, and malaria interventions, enhancing service coverage, diagnostics, and treatment adherence (Extended Data Table 1). The horizontal approach included different levels (1%, 4.85%, 6%) of annual scale-up of all HRH, targeted instantaneous scale-up of primary healthcare workforce, improvements in the availability of consumables to benchmark levels, and a comprehensive ‘HSS Expansion Package’ that combines these components alongside increased patient demand (Extended Data Table 1 and Extended Data Figure 1). The diagonal approach jointly implemented expanded HTM interventions with the full HSS expansion package. Health outcomes were measured using disability-adjusted life years (DALYs), mortality, and health service coverage.

The different investment approaches impose varying costs, making it essential to assess whether additional spending yields proportionate improvements in health outcomes - ensuring value for money. This was assessed using Incremental cost-effectiveness ratios (ICERs) relative to a common baseline scenario. ICERs represent the ratio of the incremental cost of an investment to the DALYs averted through its implementation compared to the baseline scenario or status quo. Additionally, we report the return on investment (ROI), a metric commonly used by donors^26^ which was estimated as the ratio between the net monetary benefit (calculated estimating the value that would be placed on the increases in health by the persons affected, minus the incremental cost) and the incremental cost ^26–28^.

## Results

With no expansion in health system capacity or disease program coverage above 2024 levels, we project 2.13 million deaths (95th percentile uncertainty interval [UI] 2.11 - 2.17 million) and 139.46 million DALYs (UI: 137.81 - 141.42 million) would occur between 2025-2035. We present comparative findings for the horizontal investments (HSS expansion package) relative to the joint HTM program below, as well as the combined diagonal investment (HSS + HTM package). Results for the individual components of the HSS package, and for each disease-specific programme (HIV, TB, and malaria) alone and in combination with HSS components, are also shown in Figure 1.

**Figure 1.**
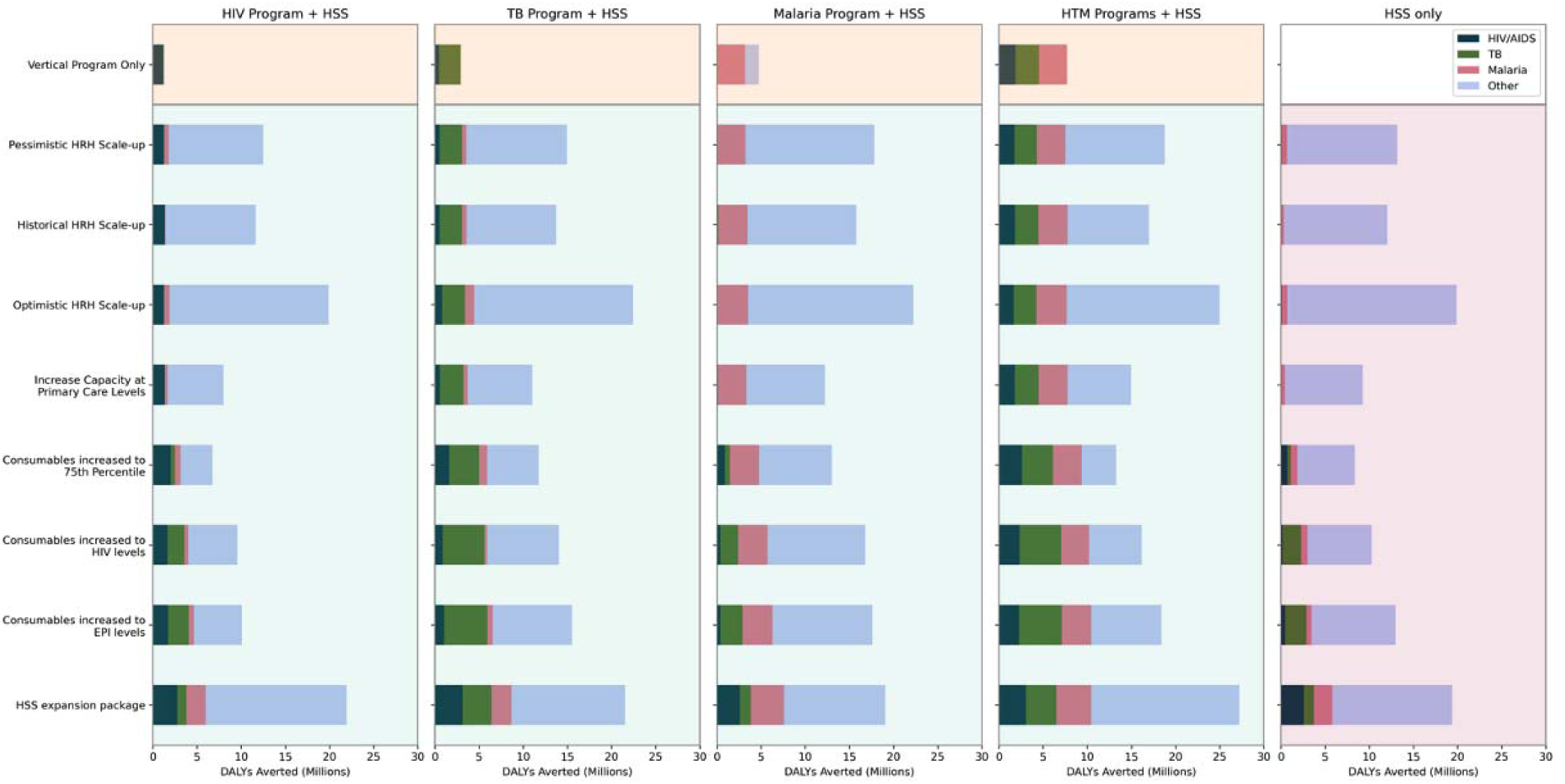
Disability-Adjusted Life Years (DALYs) averted (in millions) from 2025 to 2035, disaggregated by cause (HIV/AIDS, tuberculosis (TB), malaria, and “Other” – encompassing all remaining causes), for each intervention scenario. The shaded areas represent the three investment strategies: vertical (orange), horizontal (red) and diagonal (green). The comparison is against the *Baseline* scenario, in which health system capacity and service delivery remain fixed at 2024 levels throughout the projection period. Undiscounted values are presented. Panels 1–3 illustrate the effects of disease-program scale-up alone (Vertical Program Only - first row) and then disease program scale-up alongside each component of health system strengthening (HSS). Panel 4 shows the outputs for the joint HIV, TB and malaria (HTM) programs without and with HSS. The final panel presents the individual components of HSS. Values shown are the medians of five runs.

### Vertical investments (disease-specific health programs)

Vertical investments in HTM programs reduced disease burden but the scale of impact remained constrained by system bottlenecks that limited diagnosis, treatment availability, and service delivery (Figure 1). Jointly, the HTM programs averted 6.07 million (UI: 4.44 - 7.36 million) DALYs (UI: 4.3%, 3.2 - 5.3%) due to all causes over 11 years, reducing DALYs due to HIV, TB and malaria by 24.8% (UI: 22.3 - 26.0%), 49.7% (UI: 47.8 - 54.6%) and 64.7% (UI: 61.7 - 67.9%) respectively. This would prevent approximately 130,600 deaths due to HIV, TB and malaria. Treatment coverage rates improved for HIV and TB, from 90.2% (UI: 91.1 -– 91.5%) in 2024 to 98.1% (UI: 98.2 - 98.8%) in 2035 for HIV, from 33.8% (UI: 35.6 - 37.7%) to 59.4% (UI: 65.0 -– 67.7%) for TB, although malaria treatment rates fell from 62.6% (UI: 63.2 - 64.2%) to 55.3% (UI: 57.4 - 58.1%) due to system constraints.

### Horizontal investments (health systems strengthening)

Between 2025 and 2035, HRH investments averted 9.8% (UI: 8.6 - 10.8%), 8.6% (UI: 7.2 - 9.6%) and 14.0% (UI: 12.8 - 15.2%) of DALYs under the *Pessimistic, Historical*, and *Optimistic HRH* scale-up scenarios, respectively, compared to *Baseline*. *Primary Healthcare Workforce Scale-up* (UI: 34% immediate scale-up on primary care workforce alone) averted 6.2% (UI: 4.2 - 7.3%) of DALYs. Strengthening supply chains averted 7.6% (UI: 7.0 - 9.2%) and 9.6% (UI: 8.5 - 10.7%) of DALYs under the *Consumables increased to HIV levels* and *Consumables increased to EPI levels* scenarios respectively, with respiratory infections and diarrhoeal diseases benefiting most significantly. The *HSS expansion package* (combining *Optimistic HRH scale-up*, *Primary Healthcare Workforce Scale-up* alongside *Consumables increased to 75th percentile*) averted 18.9% (UI: 17.8 – 19.3%) of DALYs. These investments contributed to substantial mortality reductions, averting deaths due to HIV/AIDS (50,000), TB (18,300), malaria (26,800) and other causes (109,600).

### Diagonal investments (both disease programs and health system strengthening)

Diagonal approaches consistently outperformed all vertical approaches, reducing DALYs by 19.0% (UI: 17.8 - 19.8%) compared to a 4.4% reduction through vertical investments alone (Figure 1). A substantial proportion of these gains (UI: 61.5%: 26.4 million DALYs [24.5 - 28.0 million]), were for non-HTM conditions, highlighting the broader health benefits of system-wide strengthening in addressing both communicable and non-communicable diseases. Vertical programs alone could avert 4.9% (UI: 3.7 - 5.8%) of deaths, whereas diagonal investments could avert 15.8% (UI: 14.5 - 16.8%) of deaths, an additional 234,000 lives saved.

Furthermore, integrating HSS with vertical programme expansion led to greater reductions in disease burden for each of the HIV, TB, and malaria programmes (Extended Data Figure 2). Compared with vertical investments alone, this diagonal approach averted an additional 26,000 (33.3%) HIV, 15,000 (29.8%) TB, and 8,700 (43.2%) malaria deaths.

#### Value for money

The horizontal investment approaches consistently yielded greater health benefits at comparable costs relative to vertical approaches (Figure 2). Moreover, these two approaches exhibited strong synergies and economies of scale, as evidenced by the incremental health gains and cost observed when they are combined under diagonal investments. The ICER of *Joint HTM programme scale-up* without the HSS Expansion Package relative to the *Baseline* scenario was $131.62 [95% UI: $115.26 - $188.18] per DALY averted, considering only service level costs in line with standard cost-effectiveness studies^29^. The ICER of the *Joint HTM programme scale-up with HSS Expansion Package* relative to the *Baseline* scenario was $108.77 [UI: $106.19 - $122.43], demonstrating that the diagonal approach was more cost-effective than the vertical approach. While the *HSS Expansion Package* averted 10.7 [UI: 9.44 - 12.33] million more DALYs than *Joint HTM scale-up*, it did so at a higher cost per DALY averted ($143.63 [UI: $126.43 - $152.76] versus $131.62 [UI: $115.26 - $188.18]), meaning it was less cost-effective. Among the three, the diagonal approach not only achieved the greatest health impact but also offered the greatest value for money. Extended Data Table 3 provides a detailed breakdown of ICERs for all scenarios compared against the *Baseline*. The relative cost-effectiveness of scenarios remains qualitatively the same under alternative discount rate assumptions (Supplementary Figures B1 and B2).

**Figure 2.**
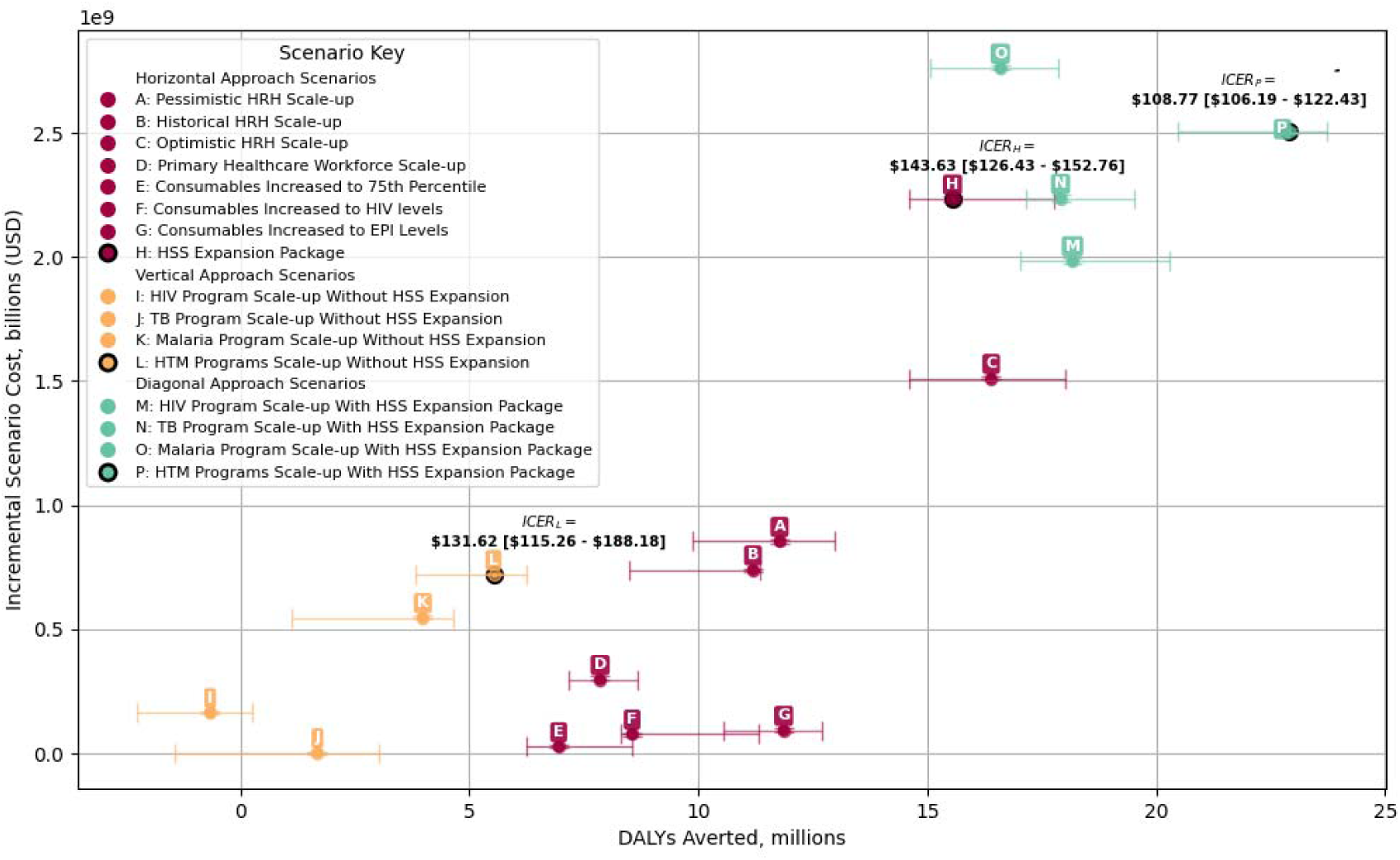
DALYs averted and incremental costs (compared to baseline) over the 11-year period (2025-2035) for all scenarios considered, categorised into horizontal, vertical and diagonal approaches. Cost and DALYs averted are both discounted annually at 3%. The origin of the plot represents the *Baseline* Scenario. The ratio of the incremental cost to the DALYs averted represents the Incremental Cost-Effectiveness Ratio of the scenarios, compared to the *Baseline* Scenario.

#### Return on investment

At a value of a statistical life year of $834^30^ and assuming no incremental above service level costs^31^, the median return on investment (ROI) for the diagonal approach was 24.94% higher than that of the *Joint HTM scale-up* approach, reaching 6.67 [UI: 5.81 - 6.85] compared to 5.34 [UI: 3.44 - 6.24] (Figure 3). Assuming no incremental costs above service level costs for the vertical approach, the diagonal approach remained more favourable provided that its incremental above service level costs did not exceed $508.72 million - equivalent to 20.29% of the diagonal approach’s incremental service-level cost - over the 2025 and 2035 period. Under an alternative assumption that the vertical approach incurs incremental above service level costs equal to 58% of its incremental service level cost (based on estimates from Opuni *et al* (2023)^31^), the diagonal approach provided a higher ROI up to an even higher threshold of $1,404.58 million incremental above service level costs in comparison with the vertical approach, or equal to 74.12% of its own incremental service level cost. These thresholds are represented by horizontal lines ‘a’ and ‘b’, respectively, in Figure 3. These results demonstrate that even with conservative assumptions around unknown implementation costs of the two approaches, concurrent investments in HSS alongside HTM programs are likely to result in a greater value for money than investing in vertical program expansion alone. Furthermore, the ROI of the diagonal approach also surpassed that of the horizontal approach (4.81 [UI: 4.46 - 5.60]). This finding remained consistent under alternate assumptions of discount rates and VSLY (Supplementary Figure B3 and B4, Supplementary Tables B1-B4).

**Figure 3.**
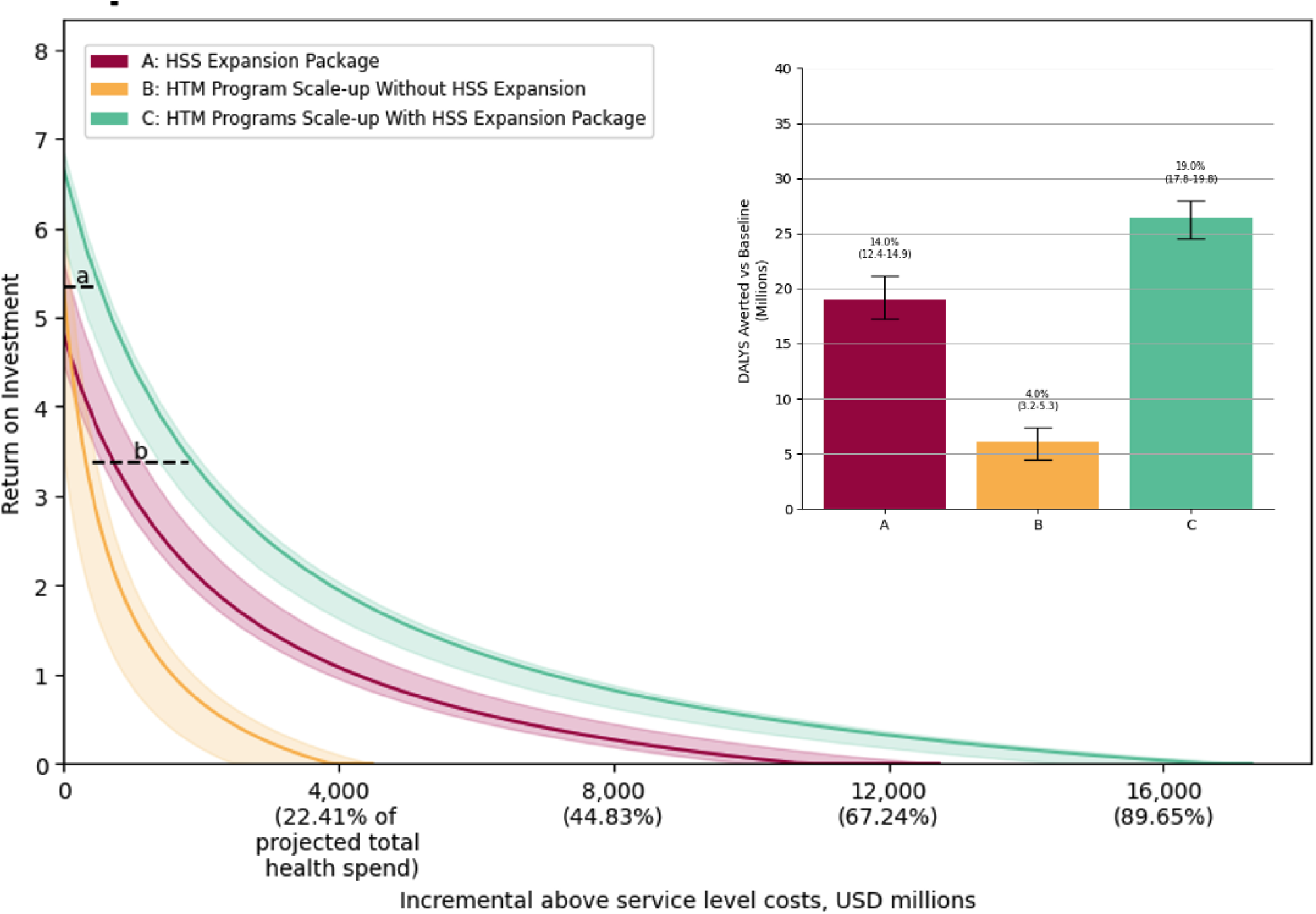
Return on investment (ROI) for (A) *HSS Expansion Package*, (B) *HTM Program Scale-up Without HSS Expansion*, and (C) *HTM Program Scale-up With HSS Expansion Package*, across a range of hypothetical incremental above service level costs. Inset: the DALYs averted relative to *Baseline* for the 3 scenarios. Percentage DALYs averted over the 11-year period compared to *Baseline* are annotated above each bar. Horizontal annotations **a** and **b** represent the maximum allowable incremental above service level cost for the diagonal scenario (in comparison with the vertical scenario) such that its ROI remains greater than that of the vertical scenario. **a** assumes zero incremental above service level costs for the vertical scenario; **b** assumes an incremental above service level costs equal to 58%^31^ of the incremental service level cost for the vertical scenario. See Results for quantitative values.

## Discussion

This study quantifies for the first time the health impact of health system strengthening, showing that integrating HSS with disease-specific program scale-up delivers substantially greater health benefits and value for money than standalone vertical interventions. The diagonal approach averted almost four times as many DALYs as vertical programs alone, primarily due to accelerated expansion of the health workforce and improved availability of essential consumables. It also proved more cost-effective, with an ICER of $109 per DALY averted against the baseline, compared to vertical HTM expansion, which had an ICER of $132. This aligns with earlier studies showing the importance of addressing system constraints to maximise the effectiveness of disease-specific interventions ^1,9,16^.

Numerous previous studies emphasize that disease-specific programs often face diminishing returns in constrained health systems^1,14–17,32^. These results support the growing body of evidence advocating for diagonal approaches in health sector investments. By improving workforce capacity and consumable availability, integrated strategies not only enhance the effectiveness of HTM programs but also strengthen the overall sustainability and effectiveness of the health system^33,34^.

While our individual-based simulation model accounts for real-world constraints such as workforce shortages and stockouts, uncertainties in epidemiological parameters and cost estimates may affect projection precision. We incorporate stochastic variation in key processes, reporting uncertainty across aggregated runs and consider a number of alternative assumptions through the sensitivity analysis. Although the *Historical HRH Scale-up* increased the health workforce more rapidly than the *Pessimistic HRH Scale-up*, it averted fewer DALYs, suggesting that other health system bottlenecks may have constrained the translation of workforce expansion into health gains; the overlap in uncertainty intervals indicates that these differences should be interpreted with caution. Similarly, despite positive health gains for HIV-specific outcomes, overall DALYs and deaths increased under the *HIV Program scale-up* without accompanying health system strengthening (-0.79% DALYs and -0.47% deaths averted). This counterintuitive result likely reflects the high resource demands of intensified HIV services (such as repeated appointments and follow-up care) outstripping available health system capacity in the absence of expanded human resources and consumables. Consequently, care for other conditions was constrained, leading to a net increase in DALYs and deaths from non-HIV causes.

Our ICER and ROI estimates reflect both direct costs of expanding health system inputs and attributable indirect costs under vertical and horizontal expansion scenarios. However, they do not account for potential additional administrative, regulatory, and implementation costs (above service-level costs^31^) that may arise when operationalizing these scenarios and so we present ROI figures across a range of hypothetical above service level costs over which the ROI remains positive. This is particularly important in the case of modelling improvements in consumable availability, for which our incremental costs include the cost of additional consumables stocked (along with proportional supply chain costs) and not the broader operational interventions that may be needed to enable these improvements.^23^. Additionally, our costing method does not systematically account for incremental equipment costs relative to the baseline^36^. This is because the quantity of medical equipment is assumed to match the levels prioritized in the national strategic plan across all scenarios, due to a lack of data on the throughput of each equipment type.

In the absence of robust estimates of such expenditures tied to specific vertical and horizontal system expansion options, we instead identify threshold levels of above service-level costs over which the diagonal approach would still yield a higher ROI than the vertical approach. The two illustrative thresholds, assuming either no incremental above service level cost for the vertical scenario or a 58% overhead based on estimates from Opuni et al. (2023), serve as lower and upper bounds for interpretation. The fact that the diagonal approach continues to outperform the vertical strategy across such a wide range of above service cost assumptions lends confidence to the robustness of our conclusions.

While our model accounts for several important constraints within the health system, there remain additional factors such as weak information systems, inadequate infrastructure, and governance challenges that could influence both the effectiveness and wider applicability of HSS efforts. In these scenarios, we have assumed an immediate rise in healthcare-seeking behaviour, modelled as almost perfect healthcare utilisation following improvements in appointment availability and workforce numbers. However, increases in demand alone are unlikely to translate into substantial health gains without parallel investments to expand the health system’s capacity^37^. Although we model the expansion of the health workforce and improvements in supply chains as mechanisms to increase service coverage, it is important to recognise that the success of these measures also depends heavily on factors such as the quality of training, staff retention, and the equitable distribution of personnel across different regions and levels of care ^38^. Strengthening health systems requires consistent, long-term investment, and the benefits of such efforts are rarely immediate. While our projections are based on relatively rapid improvements, particularly in the expansion of the health workforce, it is likely that real-world progress would unfold more gradually, shaped by local contexts and operational constraints. Although integrating vertical disease programmes with broader system-strengthening strategies can offer important synergies, the reality of limited resources means that tensions and trade-offs inevitably arise. In the simulated scenarios, services are ordered by priority with emergency care addressed first, followed by HIV, tuberculosis, malaria, and subsequently other services. Yet, even with efforts to increase HRH in key programmes, there is a risk that staff may be diverted from other essential areas of care. Careful, context-specific analysis of how resources are distributed, and the consequences of prioritisation decisions, is therefore critical to maximise impact and ensure sustainability.

The evidence from this work points to the need for a significant shift in global health financing ^1,39^. With growing financial pressures and changing donor interests, it is no longer viable to rely solely on narrowly focused, vertical programmes if meaningful improvements in health outcomes are to be achieved. What is needed are approaches that combine disease-specific priorities with efforts to tackle broader weaknesses in health systems. Such integrated strategies offer a more practical, resilient, and ultimately cost-effective route to improving health at the population level. They also help to safeguard against the uncertainties of fluctuating donor support, placing countries in a stronger position to deliver a wider range of essential services through their own health systems, with the long-term aim of building sustainability.

## Methods

### Study design and model overview

The modelling analysis was performed using the Thanzi La Onse (TLO) model, an individual-based simulation, to estimate the health impact of investments in HRH, consumables, and disease-specific (HTM) programs in Malawi^24^. The model represents interactions between individuals and the health system, capturing a wide spectrum of conditions (e.g. communicable diseases, non-communicable diseases, maternal and newborn health) and incorporating demographic, behavioural, and epidemiological processes. Detailed model structure and calibration procedures are described elsewhere ^24,40^.

Healthcare interactions are modelled as discrete, timelJordered events that capture the full complexity of service delivery within a constrained health system. Each interaction is operationalised through a detailed ‘appointment footprint’ that quantifies the clinical, nursing, pharmacy, and radiography resources (personnel, patient-facing time, consumables and equipment) required for a given service. This approach not only defines the resource use associated with routine care but also dynamically accounts for disruptions due to consumable or health workforce shortages; when appointments cannot run, protocollJspecific repeat appointments and/or referral pathways to higher-level facilities will occur.

### Health system structure

Malawi’s health system is structured into three levels: primary, secondary, and tertiary care, each serving distinct roles for healthcare delivery. Primary care encompasses community-based outreach, health posts, dispensaries, urban health centres, and primary health centres (collectively referred to as level 1a in the TLO model), and rural or community hospitals (referred to as level 1b)^21^. These facilities provide outpatient, maternity, and antenatal services. Secondary care is delivered through 26 district hospitals, each offering services available at the primary level, supplemented by additional capabilities such as x-ray imaging, ambulance services, operating theatres, and laboratory facilities. Tertiary care is provided by four central hospitals located in major urban areas, offering specialised services and advanced medical care. The Ministry of Health (MoH) oversees national policy implementation, with service delivery managed at the district level through District Health Management Teams. Health facilities across all levels are required to maintain records of medical consumables through an electronic stock management tool (OpenLMIS system)^41^, which collects and aggregates data on drug stock levels, usage rates, and stockouts.

The TLO model incorporates detailed data on the healthcare workforce in Malawi to assess the alignment between healthcare service demand and workforce capacity^21^. This integration is achieved by formalising how the time of different healthcare worker cadres - including clinicians, nurses and midwives, pharmacists, and other essential staff - is allocated to healthcare services. The model utilises data from routine health management information systems and official records, incorporating only filled positions (excluding funded but vacant posts) to provide a comprehensive view of workforce capacity. We assume that the productivity of healthcare workers to deliver healthcare services remains fixed over the duration of the simulation.

The human resources data provides detailed estimates of patient-facing time available per day for different health worker cadres across each facility level in Malawi^20^. These estimates provide an empirically grounded estimate of health worker availability and were used to assess the capacity of the health system to deliver services and model the impact of workforce constraints on health outcomes.

Appointment durations for healthcare services in Malawi are determined based on estimates of patient-facing minutes required for 49 different categories of appointments^21^, considering the average duration required for consultations, diagnostics, and treatment. These estimates help assess the capacity of healthcare workers and the number of patients they can serve within a given time frame, thus informing the modelled constraints on healthcare delivery^42^.

### HIV, tuberculosis and malaria (HTM) programs

The HIV program delivers a comprehensive suite of interventions, including voluntary testing and counselling, initiation of first-line antiretroviral therapy (ART), and targeted preventive measures such as voluntary medical male circumcision (VMMC) and pre-exposure prophylaxis (PrEP) for select high-risk populations. Treatment protocols are lifelong and aimed at achieving viral suppression; poor adherence and/or stockouts in consumables can lead to individuals defaulting from treatment and re-initiating at a later stage. The TB program employs established diagnostic modalities (principally sputum smear microscopy and GeneXpert assays) to identify active TB cases, which are then managed using standardised treatment regimens for both drug-sensitive and multidrug-resistant TB, supplemented by preventive therapy for identified paediatric contacts and people living with HIV. The malaria program is based on the widespread deployment of rapid diagnostic tests (RDTs) for case identification, followed by treatment in accordance with national guidelines for uncomplicated and severe malaria, and is supported by routine vector control measures including the distribution of insecticide-treated bednets and periodic indoor residual spraying in high-risk districts.

### Investment approaches

Three investment approaches were investigated over the period 2025–2035 with multiple scenarios considered under each (see Extended Data Table 1 for details):

1. **Horizontal investment approach:** This involves investments in health system strengthening, focusing on expansion of the healthcare workforce and improving the availability of essential medical consumables with improved healthcare-seeking behaviour in response to health system expansion.
2. **Vertical investment approach:** Targeted expansions in HIV, TB, and malaria programs within existing health system constraints, including improvements in prevention, diagnostics, and treatment coverage for each disease. Demand for HTM services is increased in response to increased program coverage.
3. **Diagonal investment approach:** This approach integrates the vertical and horizontal approaches through investments in HSS alongside HTM program scale-up, leveraging system-wide improvements to maximise the effectiveness and reach of disease-specific interventions while addressing broader health system limitations.

### Increases in HRH

To model the increase in healthcare staff over time, we used a curve fitting approach to derive a scaling factor that reflects the actual growth in staff numbers reported by Malawi MoH from 2017 to 2024 (Extended Data Figure 1a). The scaling factor for each year was estimated using an exponential function:

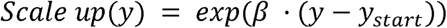

where *y* represents the year, *y_start_* is the starting year (2017), and β is a parameter defining the rate of growth. This function was fitted to historical data on HRH (using SciPy, Python v3.8) which estimates the best-fitting values for β. The data were normalised to the baseline 2017 staff number. The fitted function generated scaling factors for the years 2017 to 2024. For the years 2025-2030, the scale-up pattern was extended linearly, after which staff counts remained fixed. The average growth rate for the HRH Scale-up scenario between 2017 and 2030 was calculated using the compound annual growth rate (CAGR) formula:

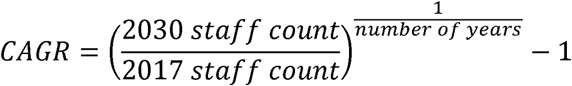

giving an average annual growth rate 4.87% from 2017 to 2030. This represents the ‘**Historical HRH Scale-up**’ scenario.

Two further generalised scenarios for HRH scale-up were considered, applying uniform annual growth rates of 6% (‘**Optimistic HRH Scale-up**’) and 1% (‘**Pessimistic HRH Scale-up**’) across all cadres and facility levels. These were implemented from 2025 to 2030. Beyond this period, we assume that workforce levels would stabilise as part of a longer-term planning framework. While the model runs through to 2035, the 2025-2030 scale-up period allows for a realistic projection of the impact of HRH investments within the typical planning and budgeting cycles used in health systems. The choice to keep workforce levels constant after 2030 is also aligned with the aim of assessing the medium-term impact of scaling up HRH under varying growth scenarios, without introducing excessive uncertainty about future workforce policies or interventions beyond the model’s timeframe.

A final HCW scenario focused specifically on scaling up only the primary healthcare workforce at facility level 1a (‘**Primary Healthcare Workforce Scale-up**’), applying the optimistic 6% growth rate. As incremental annual increases for the primary healthcare workforce alone were not operationally feasible in the model, a five-year cumulative growth of 34% was implemented as an immediate increase in 2025 to reflect a rapid workforce expansion strategy.

### Consumables availability

Three realistic scenarios were considered to modify consumable availability in Malawi at facility levels 1a and 1b, the levels where the majority of health services are delivered and where the issue of consumable stock-outs is most acute^23^. These scenarios involved adjustments to the availability of drugs, diagnostics, and other non-therapeutic supplies (e.g., gloves, syringes):

1. **Consumables Increased to 75th Percentile**: For each consumable, the availability at facility levels 1a and 1b was increased to match the benchmark facility (within the corresponding level) at the 75th percentile of availability. The final availability for each consumable was set to the higher value between its benchmark facility and the current reported availability.
2. **Consumables increased to HIV Levels**: Consumable availability was matched by facility level to the average availability of all consumables supplied through HIV program supply chains. This includes diagnostics, drugs, and HIV-specific medical items. For each consumable, availability was set to the greater of the current reported availability or the HIV program supply availability.
3. **Consumables increased to EPI Levels**: Similarly, consumable availability was matched by facility level to the average availability of all consumables supplied through the Expanded Program on Immunization (EPI) program supply chains, which included vaccines and vaccine-specific medical items (e.g. syringes, safe disposal boxes).

Extended Data Figure 1b presents the average availability of consumables by disease/public health program across these scenarios.

## Economic analysis

A Mixed-method costing approach was adopted to cost the health system resources estimated to be used under all scenarios^36^. Specifically, we applied bottom-up micro-costing to resources that are directly attributable to service delivery and apportionable by patient, such as medical consumables dispensed and health workers salaries. For higher-level costs, such as the distribution and warehousing of medical consumables (supply chain), monitoring and supervision of health workers, and other overheads, we adopted a top-down approach. The bottom-up approach uses detailed estimates of units of each resource used for healthcare delivery. The top-down approach relies on empirical estimates of the proportion of resource costs directly used for service delivery that are allocated to such overheads within Malawi. The following costs were included:

1. Medical consumables – including the cost of the quantity of consumables dispensed, quantity expected to be wasted, and supply chain overheads.
2. Human resources for health – including the cost of standard salaries, cost of in-service training and supervision overheads of the total workforce, cost of pre-service training to overcome regular health worker attrition as well as expand the workforce in certain scenarios
3. Medical equipment – including the annuitized replacement cost of equipment over their life span as well as the cost of repair, maintenance and spare parts.
4. Facility operating costs – including utility bills, maintenance, administrative staff, and other overheads.
5. Malaria scale-up costs - The full implementation cost of scaling up insecticide-treated bednet distribution and indoor residual spraying (IRS) was estimated using figures from Stelmach et al. (2018)^43^, updated with the latest consumable costs from Malawi. The cost was estimated at $1.4 per person covered by a bednet per year (assuming a 3-year lifespan and an average of 1.8 people per bednet) and $3.5 per person covered by IRS.

All costs are estimated from a healthcare provider perspective and are expressed in 2023 US dollar terms. Health benefits were quantified using Disability-Adjusted Life Years (DALYs) averted. DALYs were computed as the sum of Years of Life Lost (YLL) due to premature mortality and Years Lived with Disability (YLD)^44^. Incremental cost-effectiveness ratios (ICERs) were estimated as the ratio of incremental costs and incremental health benefit, using the following standard formula –

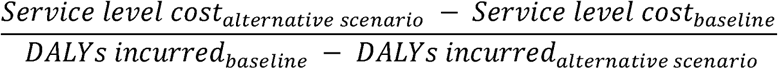

These costs represent ‘service-level costs’ which are used in standard cost-effectiveness analyses ^34^. Supplementary Table A1 presents a breakdown of incremental costs for each scenario.

To estimate Return on Investment (ROI), health benefits measured in terms of DALYs averted were monetised using an estimate of the Value of a Statistical Life Year (VSLY) in Malawi. For our main analysis, we used the VSLY estimate of $834, which has previously been used by the Government of Malawi^30^. The ROI was then estimated as follows ^26–28^

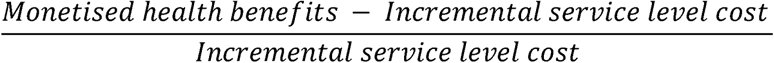

In addition to service-level costs, healthcare programs often incur substantial ‘above service level costs’, such as startup expenses, programme management, outreach, technical assistance, and broader administrative functions. For example, Opuni et al. (2023) estimated that these costs accounted for 58% of the total cost of the LINKAGES program for key populations affected by HIV in Malawi^31^. Given the absence of data on above service level costs for the scenarios modelled in this study, we report ROI estimates across a range of hypothetical values for these unknown above service level costs. To assess the robustness of our conclusions, we focus our interpretation on the range of plausible above service level costs between 0% and 58% of service-level costs. This range reflects both optimistic assumptions (i.e., minimal **incremental** above-service overheads associated with implementing each scenario) and empirical evidence from a targeted program in Malawi that is likely to have incurred relatively high above service level costs, higher than those expected under our integrated scenarios, which are designed to leverage and strengthen the existing health system infrastructure.

To incorporate above service level costs, the formula to calculate ROI is updated as follows –

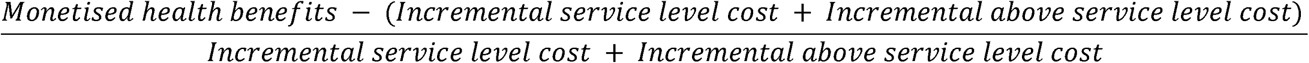

To aid the interpretation of different levels of above service level costs, we express them as a percentage of projected total health spending under the *Baseline* scenario. This is estimated by multiplying projected estimates of per capita health spending from Dieleman *et al* (2018)^45^ between 2025 and 2035 with the population projected over this period by the TLO model. Supplementary Table A2 presents the annual projected health spending between 2025 and 2035.

To avoid misleading results for scenarios where the incremental cost is negative (i.e., the scenario results in net cost savings relative to the baseline), we use in the absolute value of the incremental cost in the denominator for the ROI formula.

## Model simulations and calibration

A representative cohort (100,000 simulated individuals, i.e. a model to actual population size ratio of 1:147) of the Malawian population was simulated from 2010, assigning demographic, lifestyle and health characteristics according to reported survey, census, epidemiological and health system usage data. The model is executed in Python programming language 3.8^46^.

To account for stochastic variability, each scenario was simulated five times, and results were summarised using the median and 95% uncertainty intervals (derived from 2.5^th^ and 97.5^th^ percentiles). Comparisons across scenarios were performed using pairwise run comparisons. Stochastic variation arose from individual-level heterogeneity in disease progression, transmission dynamics, and healthcare interactions. The model was calibrated against historical epidemiological data, national databases of healthcare service delivery, including records of patient appointments, consumable stockouts, and health workforce deployment, as well as independent estimates from sources such as UNAIDS and WHO^24^.

## Sensitivity analysis

To assess the robustness of the economic conclusions, sensitivity analyses were conducted on key parameters influencing the ICERs and ROI estimates.

### Discount Rate

Health and economic outcomes were evaluated under multiple discounting assumptions to account for different time preferences regarding the value of costs and benefits, particularly relevant for interventions whose benefits accrue over the long term. Consistent with standard economic evaluations, the primary analysis applied a 3% annual discount rate to both health and costs^47^. Following WHO CHOICE methods, results were additionally reported with zero discounting of health outcome and 3% discounting of costs^48^. Finally, recognising the potential for higher growth rate in low-income countries, we reported results assuming a 5% discount rate for both health and costs^47^. Estimating ICER and ROI under varying discount rate assumptions enabled us to assess the robustness of our findings to different societal time preferences regarding the value of future costs and health benefits.

### Value of a Statistical Life Year (VSLY)

Given the inherent uncertainty in assigning a monetary value to health benefits, alternative VSLY estimates were considered. The main analysis assumed a VSLY of USD $834, reflecting prior valuation used by the Government of Malawi^30^. For our sensitivity analysis, we re-estimated the VSLY in 2025 following Robinson et al (2019)^49^, by dividing the value of a statistical life (VSL) in Malawi by the conditional life expectancy at the age equal to half of the life expectancy at birth. The VSL for Malawi was estimated using the following formula^49^,

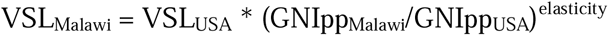

where GNIpp = Gross national income per capita, expressed in 2023 international dollars (purchasing power parity); elasticity = income elasticity of VSL

We estimate VSLY_Malawi_ as **$425.96** and **$2,427.31** in 2023 USD terms, assuming an income elasticity of VSL of 1.5^50^ and 1^49^ respectively. These provide lower and upper bound VSLY values to test the robustness of our ROI results. Evaluating ROI across this range of VSLY values was important, as lower VSLY assumptions tend to favour lower-cost interventions, even when their overall health impact is modest, while higher VSLY values, conversely, give greater weight to strategies with larger health benefits, even if their costs are higher. Supplementary Table A3 in the appendix provides details on the estimation methodology.

## Data Availability and Code Transparency

All associated code is open-source and available at https://github.com/UCL/TLOmodel/releases. Detailed model documentation, including parameter values, appointment footprints, and calibration methods, is available at Hallett *et al*^24^.

## Ethical approval

The Thanzi La Mawa project received ethical approval from the College of Medicine Malawi Research Ethics Committee (COMREC, P.09/23-0297) for the use of publicly accessible and anonymised secondary data. No data were used requiring individual informed consent.

## Extended Data

**Extended Data Table 1:**
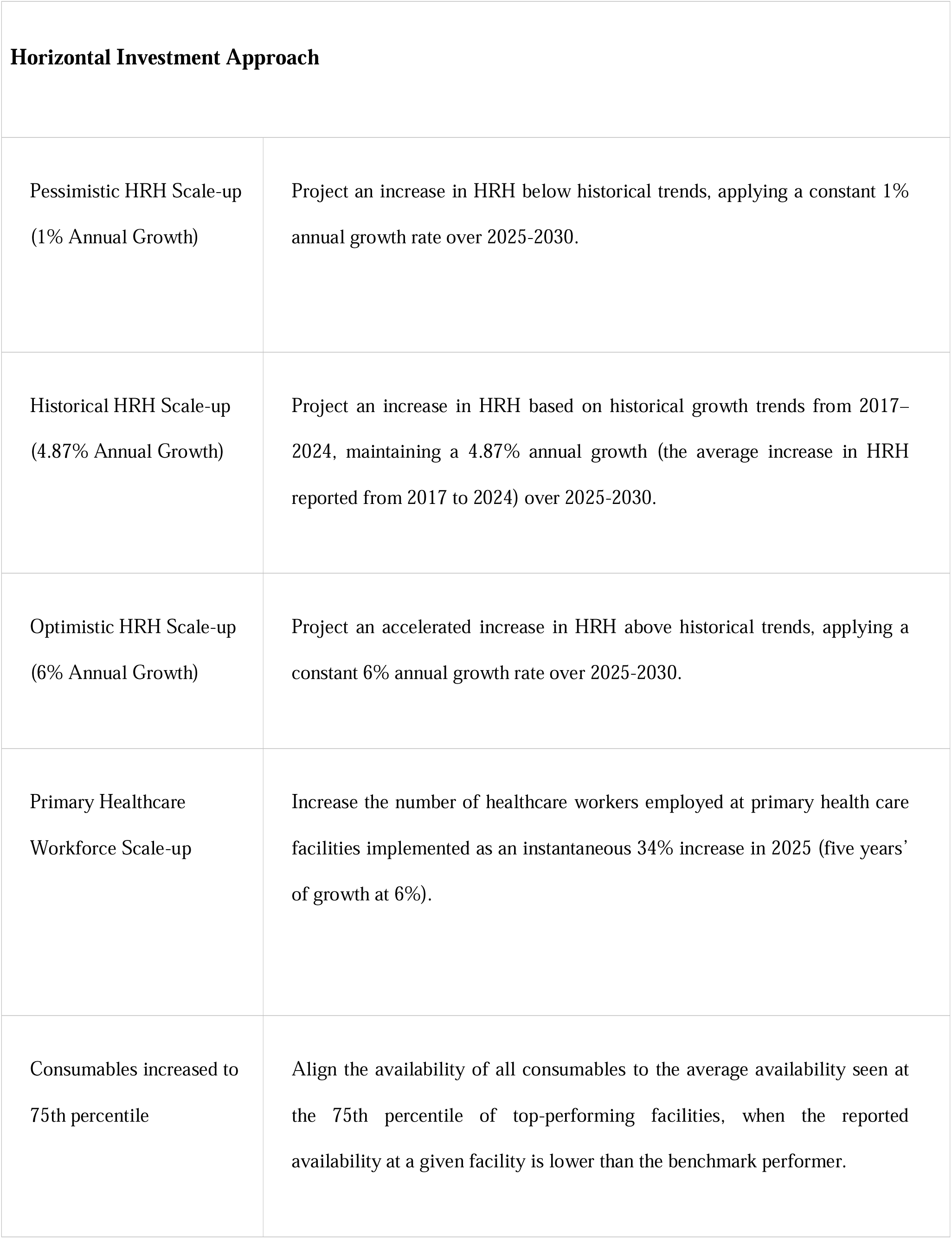
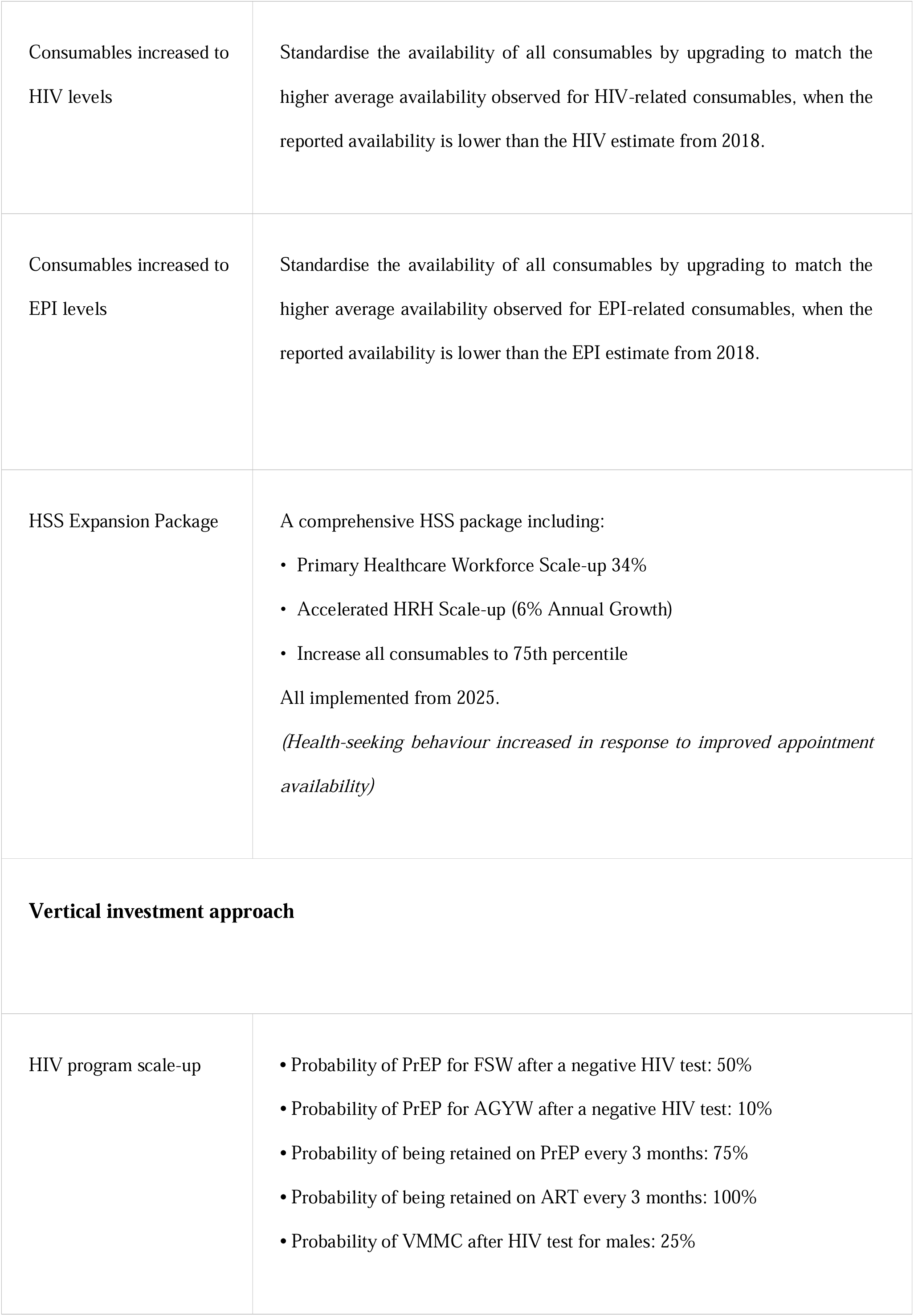
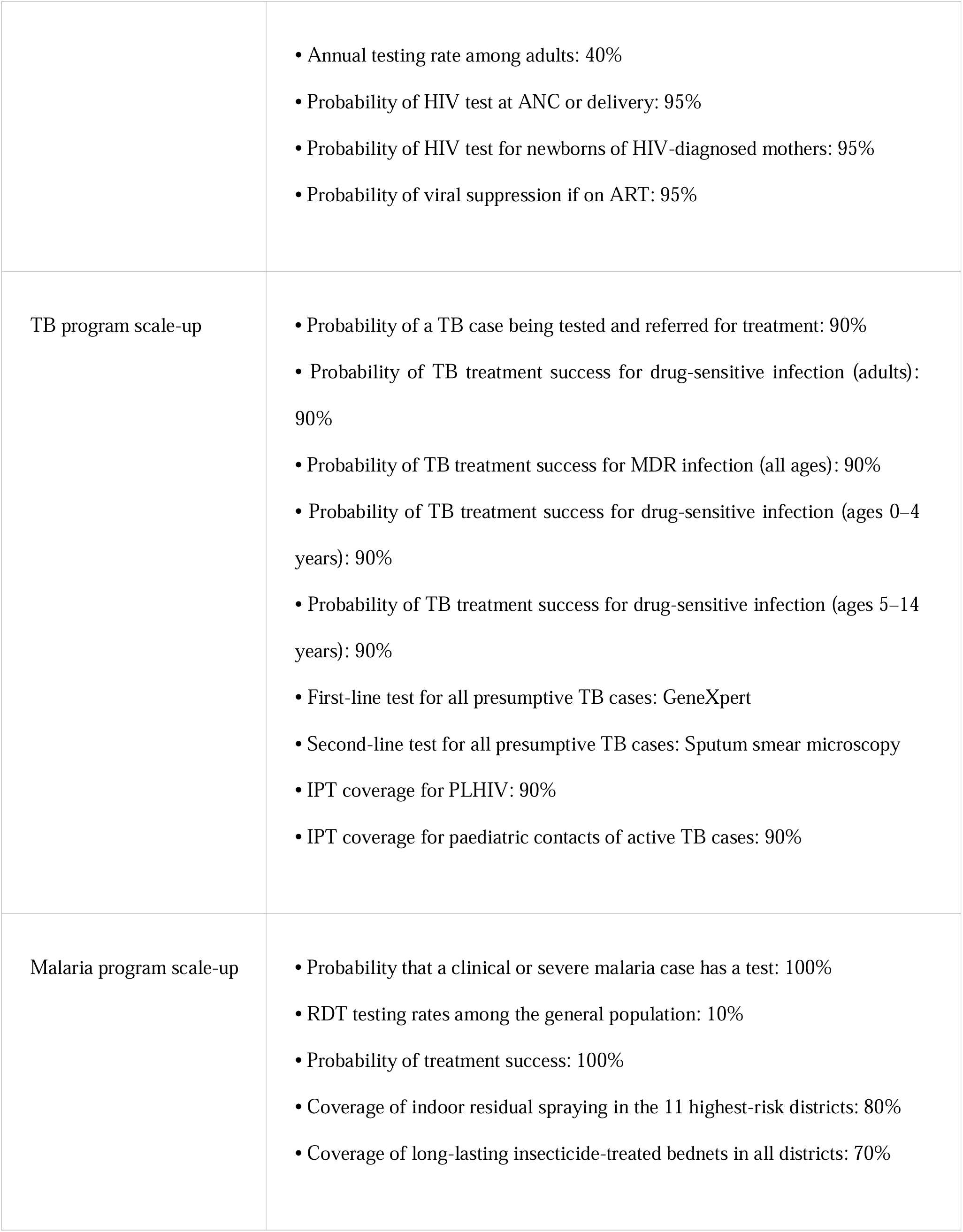
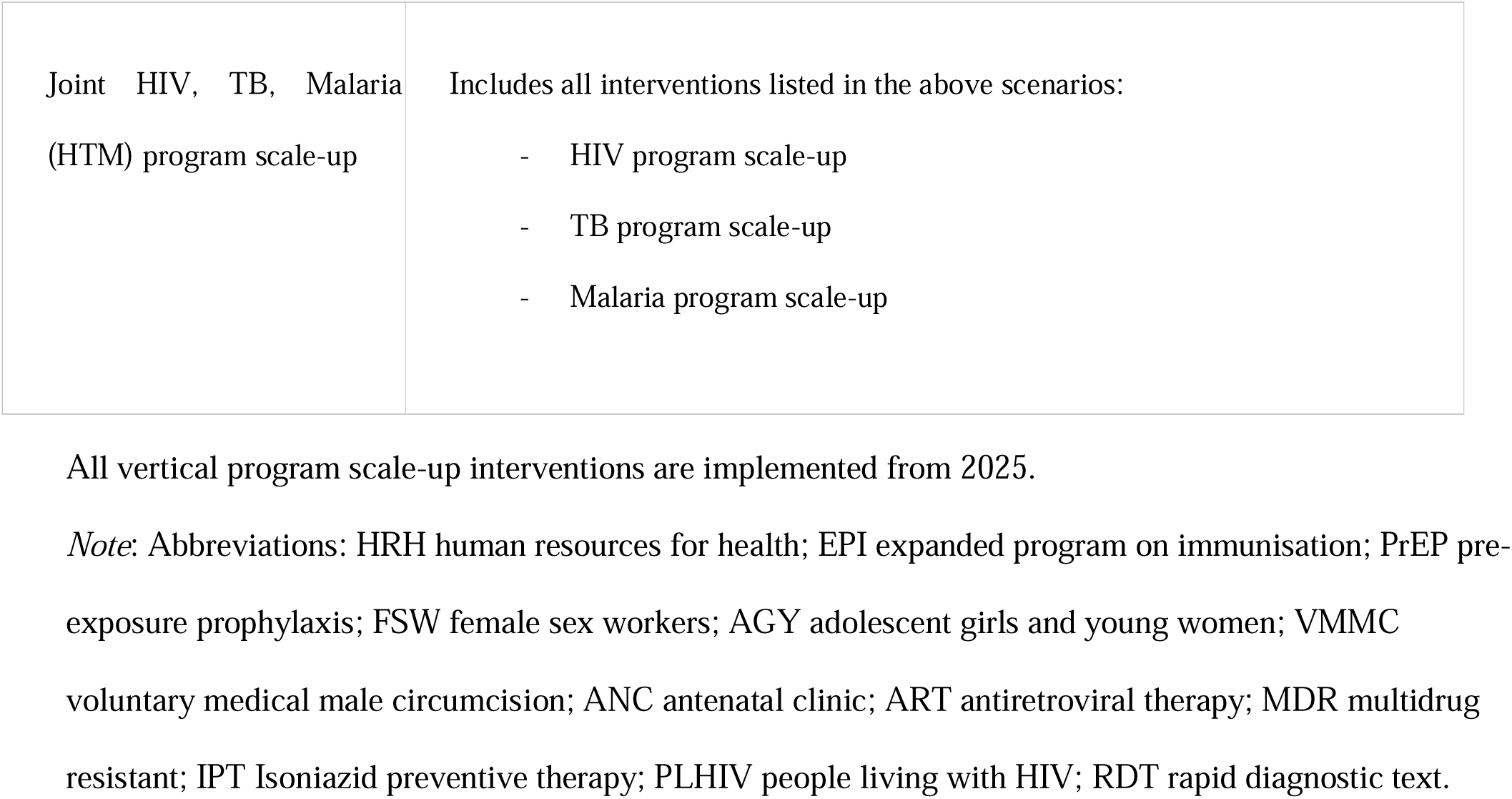
Description of scenarios.

**Extended Data Table 2:** Percentage of DALYs and deaths averted between 2025-2035 by scenario.

| **Investment approach** | **Scenario** | **Percentage DALYs averted** | **Percentage deaths averted** |
| --- | --- | --- | --- |
| **Horizontal** | Pessimistic HRH Scale-up | 9.75 (8.58 - 10.84) | 7.21 (5.91 - 8.65) |
|  | Historical HRH Scale-up | 8.63 (7.25 - 9.59) | 6.26 (5.13 - 7.15) |
|  | Optimistic HRH Scale-up | 14.02 (12.76 - 15.16) | 10.59 (9.36 - 11.75) |
|  | Primary Healthcare Workforce Scale-up | 6.24 (4.21 - 7.29) | 3.89 (1.46 - 5.22) |
|  | Consumables Increased to 75th Percentile | 6.01 (5.23 - 7.01) | 4.88 (4.10 - 5.61) |
|  | Consumables Increased to HIV levels | 7.55 (6.96 - 9.16) | 6.82 (6.10 - 8.74) |
|  | Consumables Increased to EPI levels | 9.56 (8.54 - 10.69) | 8.84 (7.86 - 9.71) |
|  | **HSS Expansion Package** | **13.63 (12.42 - 14.95)** | **10.75 (9.21 - 12.71)** |
| **Vertical** | HIV Program Scale-up Without HSS Expansion | -0.79 (-1.73 - 0.25) | -0.47 (-1.86 - 0.98) |
|  | TB Program Scale-up Without HSS Expansion | 1.04 (-1.10 - 2.37) | 1.51 (-0.12 - 2.67) |
|  | Malaria Program Scale-up Without HSS Expansion | 2.96 (1.01 - 3.98) | 2.00 (-0.01 - 2.75) |
|  | **HTM Programs Scale-up Without HSS Expansion** | **4.35 (3.19 - 5.31)** | **4.87 (3.73 - 5.75)** |
| **Diagonal** | HIV Program Scale-up With HSS Expansion Package | 15.84 (14.89 - 17.11) | 12.36 (11.19 - 13.08) |
|  | TB Program Scale-up With HSS Expansion Package | 15.58 (14.90 - 16.40) | 13.21 (12.53 - 14.59) |
|  | Malaria Program Scale-up With HSS Expansion Package | 13.96 (13.15 - 15.06) | 11.12 (9.68 - 12.19) |
|  | **HTM Programs Scale-up With HSS Expansion Package** | **18.94 (17.78 - 19.83)** | **15.76 (14.53 - 16.84)** |

Values show the median DALYs averted compared with *Baseline* across the five simulated runs for each scenario, with 95% uncertainty intervals in brackets defined as the 2.5th and 97.5th percentiles.

**Extended Data Table 3:** Incremental Cost-Effectiveness Ratio by scenario.

| **Investment approach** | **Scenario** | **ICER (2023 USD)**<br>*(in comparison with the baseline scenario which assumes that health system capacity remains at 2024 levels)* |
| --- | --- | --- |
| **Horizontal** | Pessimistic HRH Scale-up | \$71.95 [\$65.56 - \$87.17] |
| | Historical HRH Scale-up | \$65.76 [\$64.77 - \$87.25] |
| | Optimistic HRH Scale-up | \$92.01 [\$84.38 - \$103.97] |
| | Primary Healthcare Workforce Scale-up | \$39.97 [\$34.05 - \$41.91] |
| | Consumables Increased to 75th Percentile | \$3.89 [\$2.77 - \$5.45] |
| | Consumables Increased to HIV levels | \$8.91 [\$6.81 - \$9.44] |
| | Consumables Increased to EPI levels | \$8.44 [\$7.26 - \$8.78] |
| | **HSS Expansion Package** | **\$143.63 [\$126.43 - \$152.76]** |
| **Vertical** | HIV Program Scale-up Without HSS Expansion | Dominated |
| | TB Program Scale-up Without HSS Expansion | \$0.74 [\$-1.83 - \$6.04] |
| | Malaria Program Scale-up Without HSS Expansion | \$137.38 [\$117.33 - \$611.32] |
| | **HTM Programs Scale-up Without HSS Expansion** | **\$131.62 [\$115.26 - \$188.18]** |
| **Diagonal** | HIV Program Scale-up With HSS Expansion Package | \$109.55 [\$97.23 - \$116.63] |
| | TB Program Scale-up With HSS Expansion Package | \$124.79 [\$115.24 - \$130.37] |
| | Malaria Program Scale-up With HSS Expansion Package | \$167.30 [\$154.17 - \$183.42] |
| | **HTM Programs Scale-up With HSS Expansion Package** | **\$108.77 [\$106.19 - \$122.43]** |

ICER estimates only include service-level costs and assume zero above service level costs.

**Extended Data Table 4:**
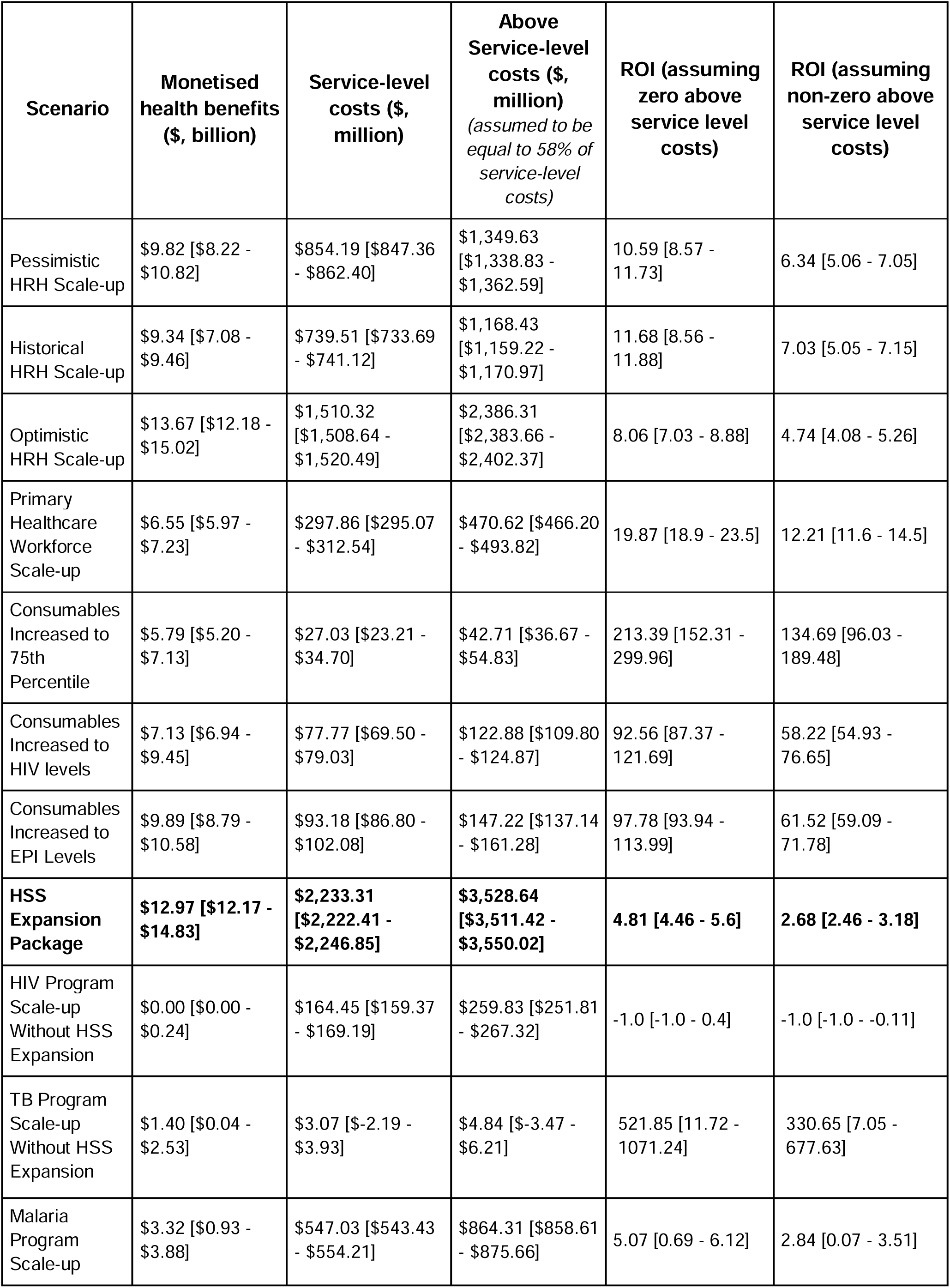
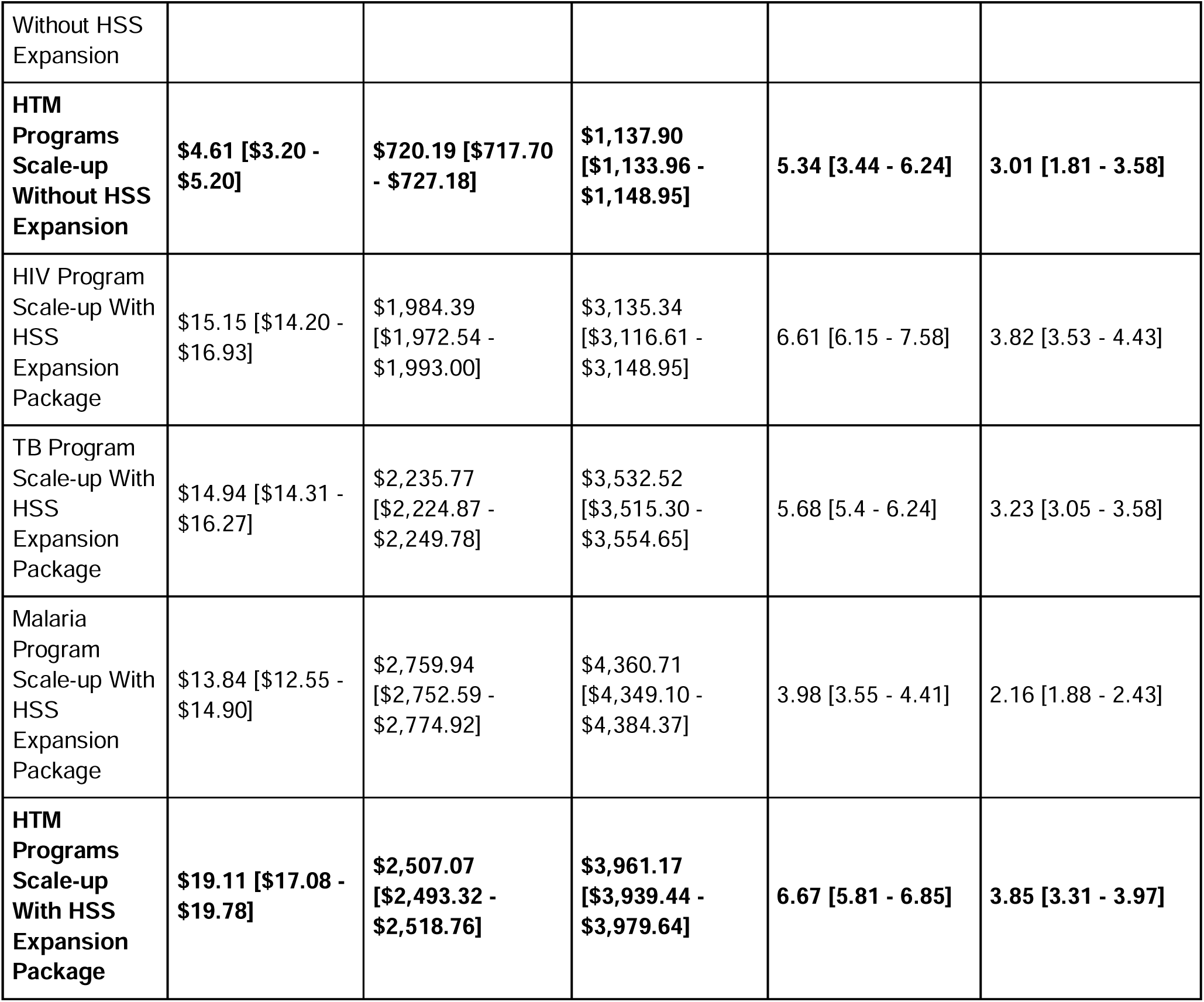
Return on Investment by Scenario (Value of Statistical Life Year = $834)

**Extended Data Figure 1.**
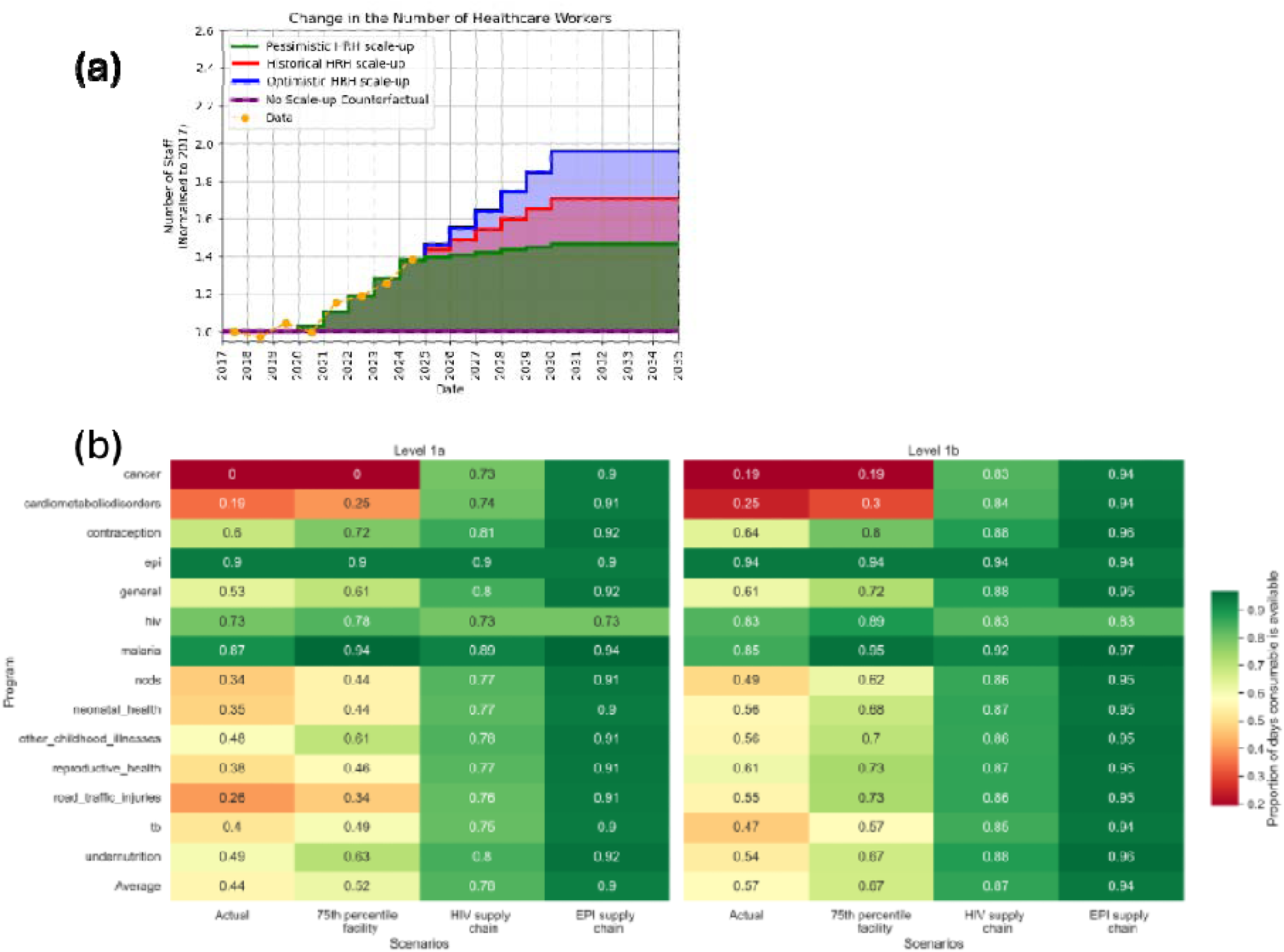
The proportional increase in the number of health workers compared to 2017 levels under 4 scenarios (a); the average availability of consumables for facility levels 1a and 1b categorised by disease/public health program under 4 scenarios – Actual (used in the Baseline scenario), Increase all consumables to 75th percentile, Consumables available at HIV levels, Consumables available at EPI levels (b).

**Extended Data Figure 2.**
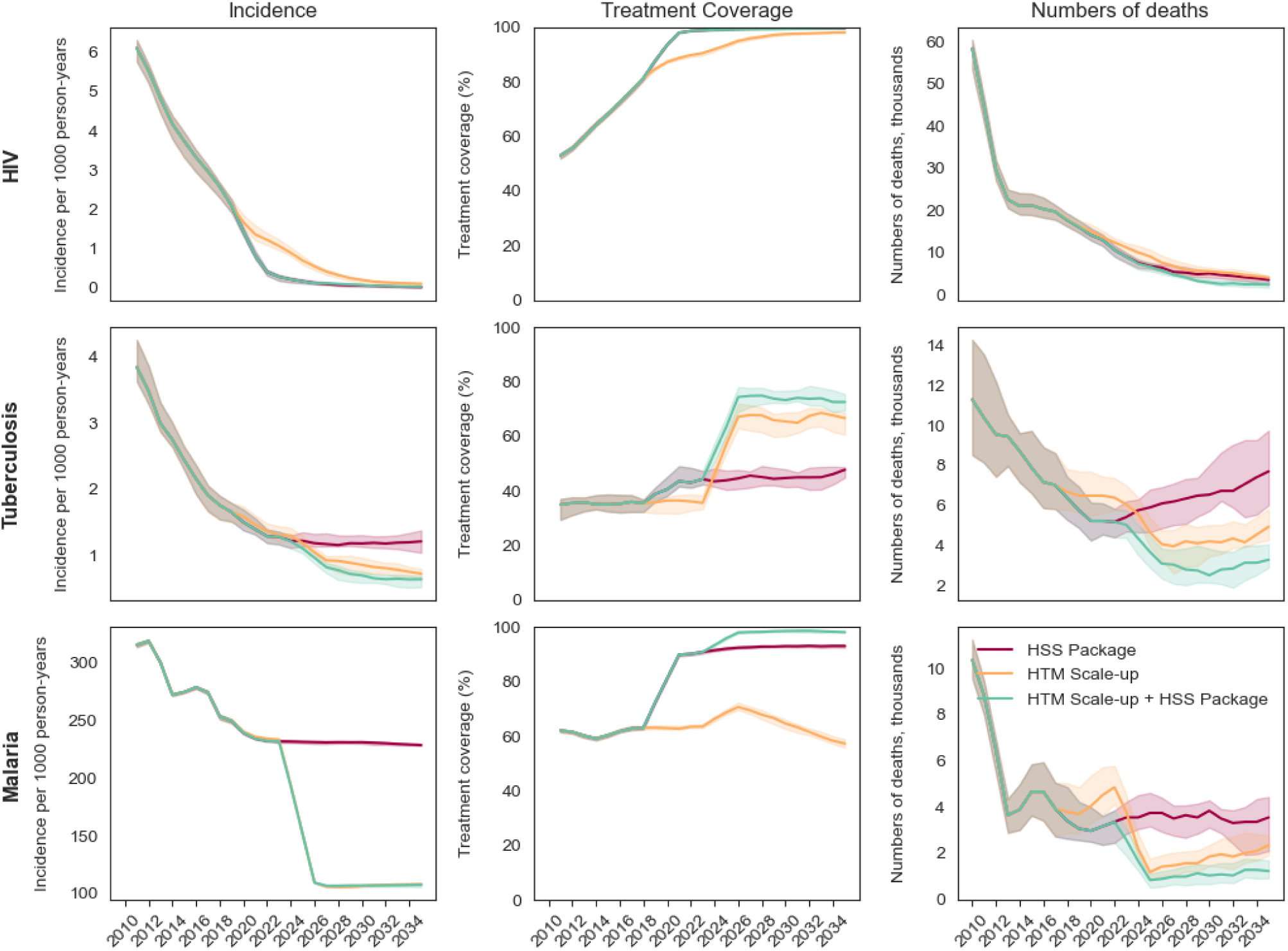
The incidence per 1000 person-years (first column), the treatment coverage (second column) and the numbers of death (third column) for each disease (HIV first row, tuberculosis second row and malaria third row) for three scenarios: HSS Expansion Package, HTM Program Scale-up, and HTM Program Scale-up with HSS Expansion Package. Outputs are shown for each year of the simulation (2010 - 2035) wi solid lines depicting the medians across 5 runs and shaded areas showing the uncertainty intervals (2.5th and 97.5th percentiles of the aggregated runs).

