## Appendix for "System-Wide Investments Enhance HIV, TB and Malaria Control in Malawi and Deliver Greater Health Impact"

A. **Economic analysis**

1. **Incremental costs**

We estimated incremental health system costs for each investment scenario across five cost categories: human resources for health, medical consumables, medical equipment, facility operations, and malaria scale-up. The costing methodology for the first four categories is detailed in Mohan et al. (2025)^1^, based on the latest available local cost data for Malawi. The costs associated with malaria scale-up—including insecticide-treated bednet distribution and indoor residual spraying (IRS)—were estimated using figures from Stelmach et al. (2018)^2^, updated with recent consumable price data from Malawi. Specifically, we assumed a cost of $1.40 per person covered by a bednet per year (based on a 3-year lifespan and an average of 1.8 people per bednet) and $3.50 per person covered by IRS annually. Supplementary Table A.1 presents a breakdown of the median incremental costs across these categories for each scenario, relative to the Baseline.

**Supplementary Table A.1: Median incremental health system cost by scenario (2025-2035)**

| **Scenario** | **Human resources for health** | **Medical consumables** | **Medical equipment** | **Facility operations** | **Malaria scale-up** | **Total** |
| --- | --- | --- | --- | --- | --- | --- |
| Pessimistic HRH Scale-up | 716,409,160 | 138,103,943 | - | - | - | 854,513,103 |
| Historical HRH Scale-up | 614,238,078 | 123,336,613 | - | - | - | 737,574,691 |
| Optimistic HRH Scale-up | 1,296,332,804 | 215,802,789 | - | - | - | 1,512,135,593 |
| Primary Healthcare Workforce Scale-up | 246,631,545 | 54,204,452 | (82) | - | - | 300,835,915 |
| Consumables Increased to 75th Percentile | - | 28,267,686 | - | - | - | 28,267,686 |
| Consumables Increased to HIV levels | - | 74,208,156 | 60,958 | - | - | 74,269,114 |
| Consumables Increased to EPI levels | - | 90,630,380 | 60,958 | - | - | 90,691,338 |
| HSS Expansion Package | 1,740,848,854 | 493,466,991 | - | - | - | 2,234,315,846 |
| HIV Program Scale-up Without HSS Expansion | - | 164,369,849 | - | - | - | 164,369,849 |
| HIV Program Scale-up With HSS Expansion Package | 1,740,848,854 | 244,829,641 | - | - | - | 1,985,678,495 |
| TB Program Scale-up Without HSS Expansion | - | 2,186,642 | - | - | - | 2,186,642 |
| TB Program Scale-up With HSS Expansion Package | 1,740,848,854 | 497,259,568 | - | - | - | 2,238,108,422 |
| Malaria Program Scale-up Without HSS Expansion | - | 2,172,274 | - | - | 545,867,917 | 548,040,191 |
| Malaria Program Scale-up With HSS Expansion Package | 1,740,848,854 | 488,938,307 | - | - | 533,086,458 | 2,762,873,619 |
| HTM Programs Scale-up Without HSS Expansion | - | 173,131,299 | - | - | 549,517,911 | 722,649,210 |
| HTM Programs Scale-up With HSS Expansion Package | 1,740,848,854 | 237,071,587 | - | - | 530,279,597 | 2,508,200,038 |

**Projected health spending**

Projected total health spending under the Baseline scenario was estimated to contextualize the scale of incremental above service level costs.

To obtain these, per capita health spending projections from Dieleman et al. (2018)^3^ were multiplied with annual population projections generated by the Thanzi La Onse (TLO) model (under the baseline scenario) for the years 2025–2035. The per capita health spending projections include health spending from all sources - government, development assistance, prepaid private spending (insurance), and out of pocket spending. Supplementary table A2 presents estimates of projected health spending for each year between 2025 and 2035 along with the discounted cumulative health spending over the full projection period, applying a 3% annual discount rate to align with our economic evaluation estimates.

**Supplementary Table A.2: Projections of health spending (Baseline scenario)**

| **Year** | **Health spending per capita (2023 USD)** | **Projected population**  **(median value)** | **Projected total health spending** |
| --- | --- | --- | --- |
|  | (A) | (B) | (A) X (B) |
| 2025 | 66.58 | 22,696,330 | 1,511,130,388.44 |
| 2026 | 67.25 | 23,464,748 | 1,577,914,897.60 |
| 2027 | 67.92 | 24,264,136 | 1,647,987,381.57 |
| 2028 | 68.60 | 25,120,519 | 1,723,213,275.86 |
| 2029 | 69.28 | 25,996,385 | 1,801,128,751.16 |
| 2030 | 69.98 | 26,858,002 | 1,879,433,130.11 |
| 2031 | 70.68 | 27,797,261 | 1,964,610,907.98 |
| 2032 | 71.38 | 28,730,413 | 2,050,868,386.98 |
| 2033 | 72.10 | 29,705,875 | 2,141,704,998.18 |
| 2034 | 72.82 | 30,686,136 | 2,234,502,640.64 |
| 2035 | 73.55 | 31,694,312 | 2,330,995,183.88 |
| Grand total projected health spending (discounted at 3%) | | | 17,846,737,270.39 |

**3. Value of a Statistical Life Year (VSLY)**

Our Return on Investment (ROI) calculations rely upon estimates of the Value of a Statistical Life Year (VSLY) to monetise the health impact of scenarios, measured in terms of Disability-adjusted Life years (DALYs) lost averted. Our main results use the VSLY figure of $834 used by the Government of Malawi^4^. Given the inherent uncertainty in assigning a monetary value to health benefits, alternative VSLY estimates were considered. These were estimated using the methodology outlined by Robinson et al. (2019)^5^. Supplementary Table A3 describes the formulae used to estimate the alternative VSLY estimates used in our sensitivity analysis. Our estimates apply two alternative assumptions for the income elasticity of the Value of a Statistical Life (VSL)—1.0 and 1.5—reflecting guidance from previous literature ^5,6^. These estimates provide us an upper and lower limit for VSLY around value of $834 used for our main results. Gross national income per capita (GNIpp) figures were sourced from World Development Indicators (2023) for Malawi and the United States, with adjustments made for purchasing power parity and currency conversions to 2023 USD. Conditional life expectancy at age 30–34 in 2025 was used to convert VSL estimates into VSLY estimates. Life expectancy was estimated using the TLO model *Baseline* scenario, extracting the age-specific all-cause mortality rates for that year and using standard life tables to estimate the life expectancy conditional on survival by age^7^.

Evaluating ROI across this range of VSLY values was important, as lower VSLY assumptions tend to favour lower-cost interventions, even when their overall health impact is modest, while higher VSLY values, conversely, give greater weight to strategies with larger health benefits, even if their costs are higher. Supplementary Table A3 in the appendix provides details on the estimation methodology.

**Supplementary Table A.3: Estimation of Value of Statistical Life Year (VSLY) For Malawi**

| **Indicator** | **Value** | **Unit** | **Source** |
| --- | --- | --- | --- |
| GNIpp_USA_ | 57,800 | 2017 PPP Dollars | Robinson et al (2020) |
| VSL_USA_ | 9,400,000 | 2017 PPP Dollars | Robinson et al (2020) |
| GNIpp_Malawi_ | 1,780 | 2023 PPP Dollars | World Development Indicators (2023) |
| LE_0_ | 62.8 | years | TLO Model projection (Baseline scenario) |
| LE_30-34_ | 40.2 | years | TLO Model projection (Baseline scenario) |
| VSL_Malawi\|elasticity = 1_ | 289,481 | 2023 US Dollars | ${(VSL}_{USA}* (\frac{{GNIpp}_{Malawi}}{{GNIpp}_{USA}})^{elasticity})*conversion factor$ |
| VSL_Malawi\|elasticity = 1.5_ | 50,800.28 | 2023 US Dollars | ${(VSL}_{USA}* (\frac{{GNIpp}_{Malawi}}{{GNIpp}_{USA}})^{elasticity})*conversion factor$ |
| **VSLY_Malawi\|elasticity = 1_** | **2427.31** | 2023 US Dollars | $\frac{\boldsymbol{VS}\boldsymbol{L}_{\boldsymbol{Malawi\vert elastcity = 1}}}{\boldsymbol{L}\boldsymbol{E}_{\boldsymbol{30-34}}}$ |
| **VSLY_Malawi\|elasticity = 1.5_** | **425.96** | 2023 US Dollars | $\frac{\boldsymbol{VS}\boldsymbol{L}_{\boldsymbol{Malawi\vert elastcity = 1.5}}}{\boldsymbol{L}\boldsymbol{E}_{\boldsymbol{30-34}}}$ |
| *Key: GNIpp = Gross national income per capita; PPP = Purchasing power parity; VSL = Value of Statistical Life; VSLY = Value of Statistical Life Year; LE_0_ = Life expectancy at birth; LE_30-34_ = Conditional life expectancy at 30-34 years of age; conversion factor = 2023 PPP dollars to 2023 US Dollars conversion rate (assumed to be 0.3371); elasticity = income elasticity of VSL*  *Sources: Robinson et al (2020)**^5^**, World Development Indicators (2023)**^8^* | | | |

**B. Sensitivity analysis**

1. **Alternative discount rates**

Health and economic outcomes were evaluated under multiple discounting assumptions to account for different time preferences regarding the value of costs and benefits. This can affect the valuation of scenarios whose health impact and cost are not evenly distributed over time. For example, a high discount rate would reduce the value of scenarios for which larger health gains are made during future years. Consistent with standard economic evaluations, the primary analysis applied a 3% annual discount rate to both health and costs^9^. Following WHO CHOICE methods, results were additionally reported with zero discounting of health outcome and 3% discounting of costs^10^. Finally, recognising the potential for higher growth rate in low-income countries, we reported results assuming a 5% discount rate for both health and costs^9^. Estimating ICER and ROI under varying discount rate assumptions enabled us to assess the robustness of our findings to different societal time preferences regarding the value of future costs and health benefits.

Supplementary Figures B1 and B2 present the cost-effectiveness plane plots of incremental costs versus DALYs averted across all scenarios under alternative discount rate assumptions. Supplementary Figure B1 applies a 0% discount rate for health outcomes and a 3% discount rate for cost, while Supplementary Figure B2 applies a 5% discount rate for both health outcomes and costs. These alternative discounting assumptions allow us to test the sensitivity of our findings to different valuations of future health benefits and costs. Across both alternative discounting scenarios, the relative cost-effectiveness ranking of the different investment strategies remains qualitatively unchanged from our main results, supporting the robustness of our main conclusions.

Supplementary Figure B3 and Supplementary Tables B1 and B2 present ROI estimates for the joint HTM scenario under the three approaches assuming alternative discount rates and a VSLY of $834. In the first case (0% discount rate for health and 3% discount rate for cost), under the assumption of zero above service level costs for the vertical approach, the ROI of the diagonal approach remains higher than the vertical approach even if its incremental above service level costs are up to $559.3 million (22.31% of the incremental service level cost of the diagonal approach or 3.13% of the total project health spending for 2025-2035). If we assume that above service level costs of the vertical approach amounting to 58% of its service level cost, the diagonal approach provided a higher ROI up to an even higher threshold of $1,567.94 million incremental above service level costs in comparison with the vertical approach (62.54% of its own incremental service level cost or 8.79% of the total project health spending for 2025-2035).

In the second case (5% discount rate for health and cost), the threshold incremental above service level costs for the diagonal approach to provide a greater ROI than the vertical approach under the two above service level cost assumptions were $446.9 (19.62% of its own incremental service level cost or 2.5% of the total project health spending for 2025-2035) and $1282.63 million (56.3% of its own incremental service level cost or 7.18% of the total project health spending for 2025-2035).

**Supplementary Figure B.1: DALYs averted and incremental costs (compared to baseline) over the 11-year period (2025-2035)** *(0% discount rate for health and 3% discount rate for costs)*

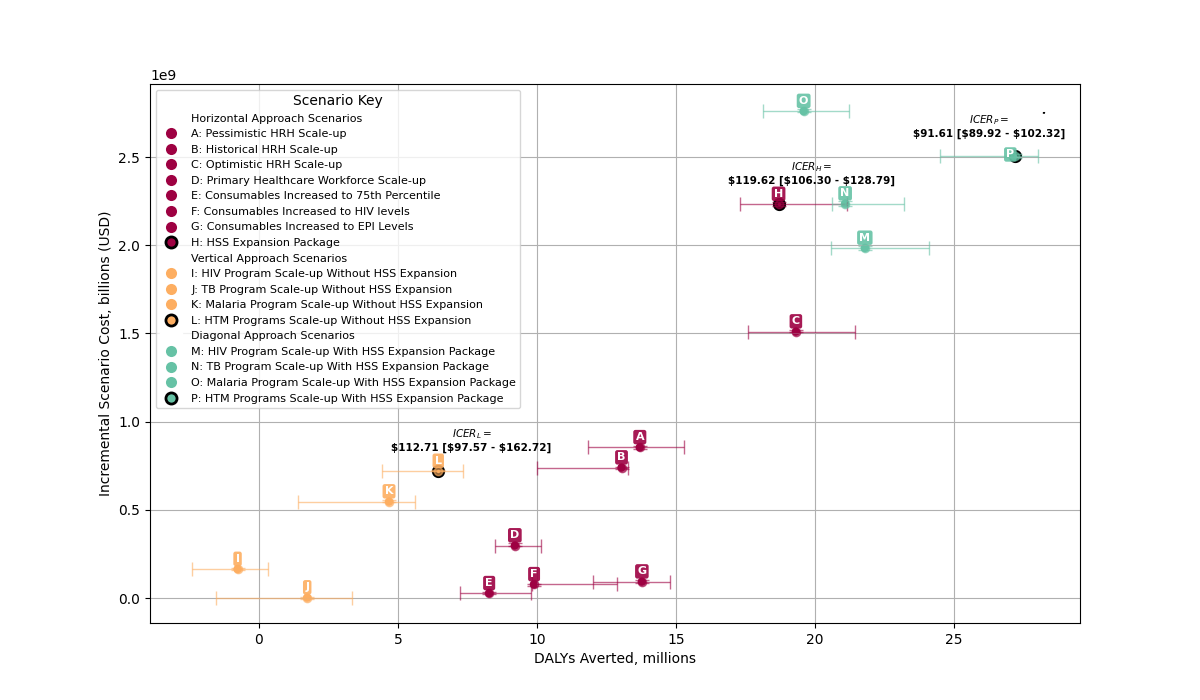

**Supplementary Figure B.2: DALYs averted and incremental costs (compared to baseline) over the 11-year period (2025-2035)** *(5% discount rate for both health and cost)*

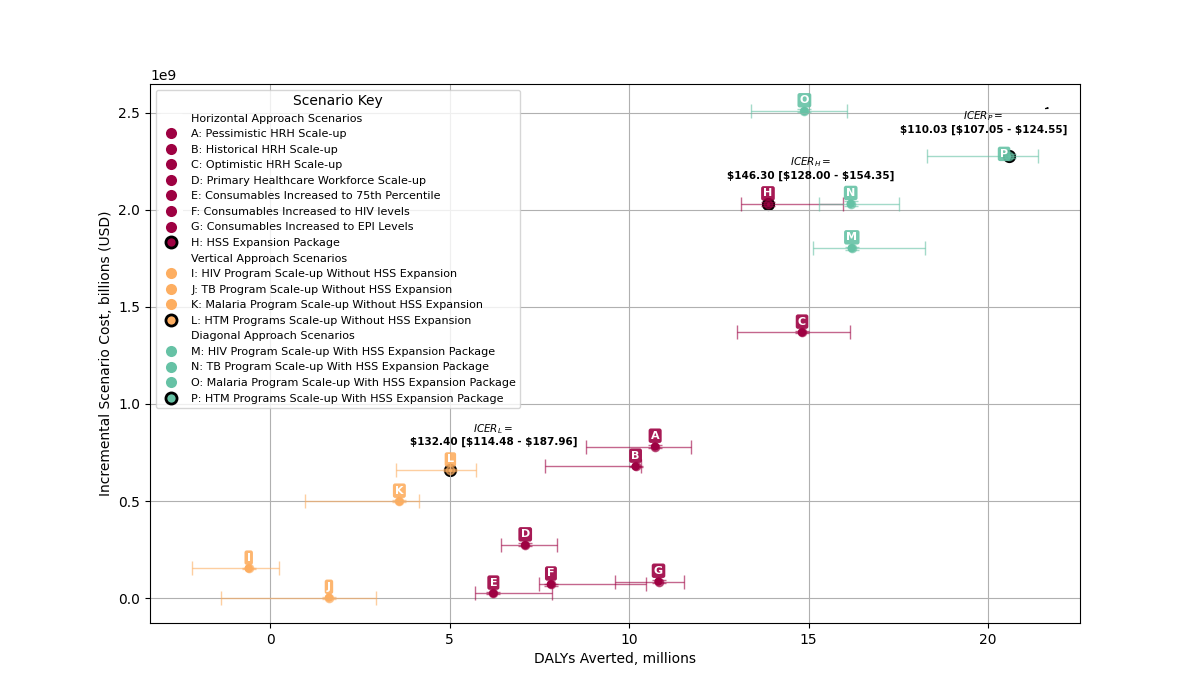

**Supplementary Figure B.3 Return on Investment (ROI) assuming alternative discount rates**

| 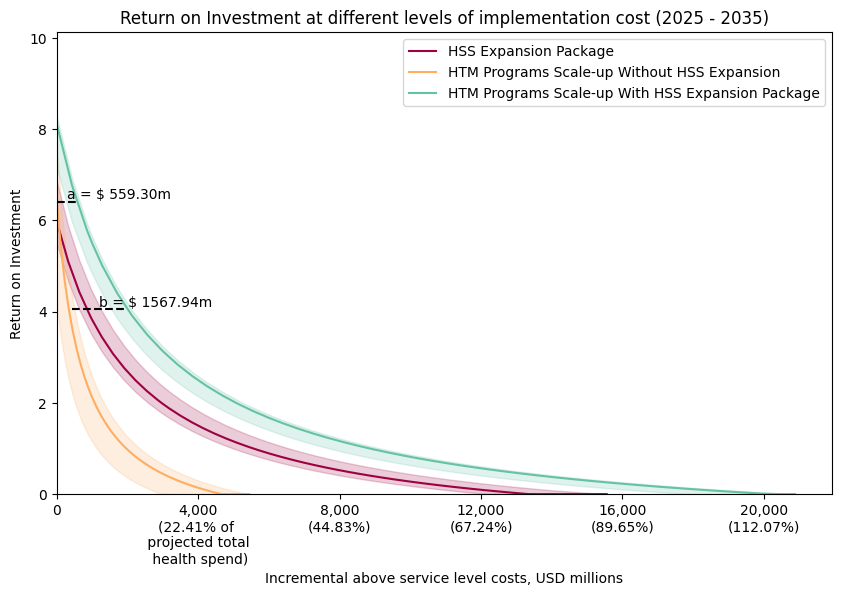 |
| --- |
| (i) 0% discount rate for health; 3% discount rate for costs  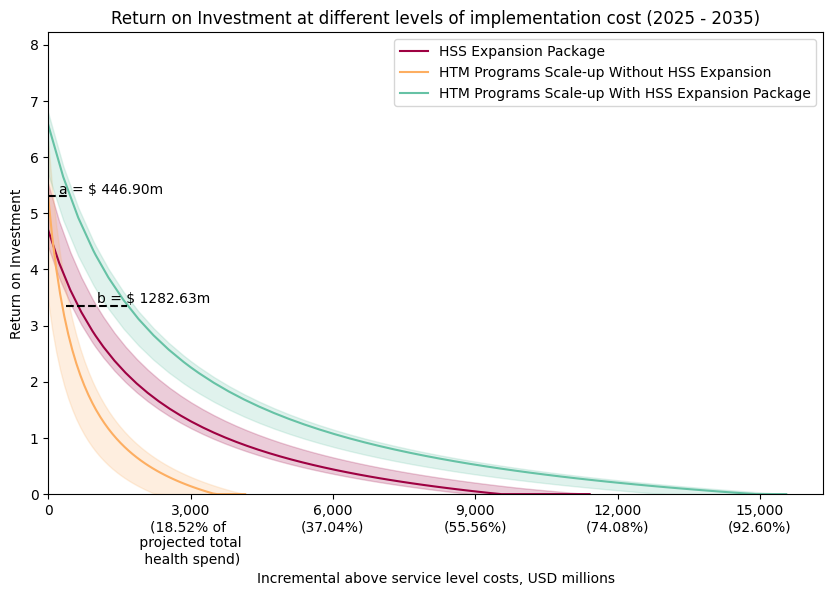  (ii) 5% discount rate for health; 5% discount rate for costs |
| * Note: ROI estimates are clipped at a minimum value of 0. VSLY is assumed to be $834. |

**Supplementary Table B.1: Return on Investment by Scenario (Value of Statistical Life Year = $834; Health discount rate = 0% and cost discount rate = 3%)**

| **Scenario** | **Monetised health benefits ($, billion)** | **Service-level costs ($, million)** | **Above Service-level costs ($, million)**  *(assumed to be equal to 58% of service-level costs)* | **ROI (assuming zero above service level costs)** | **ROI (assuming non-zero above service level costs)** |
| --- | --- | --- | --- | --- | --- |
| Pessimistic HRH Scale-up | $11.43 [$9.87 - $12.75] | $854.19 [$847.36 - $862.40] | $1,349.63 [$1,338.83 - $1,362.59] | 12.49 [10.49 - 13.98] | 7.54 [6.27 - 8.48] |
| Historical HRH Scale-up | $10.88 [$8.35 - $11.07] | $739.51 [$733.69 - $741.12] | $1,168.43 [$1,159.22 - $1,170.97] | 13.83 [10.28 - 14.0] | 8.39 [6.14 - 8.49] |
| Optimistic HRH Scale-up | $16.11 [$14.67 - $17.87] | $1,510.32 [$1,508.64 - $1,520.49] | $2,386.31 [$2,383.66 - $2,402.37] | 9.68 [8.67 - 10.76] | 5.76 [5.12 - 6.44] |
| Primary Healthcare Workforce Scale-up | $7.68 [$7.09 - $8.46] | $297.86 [$295.07 - $312.54] | $470.62 [$466.20 - $493.82] | 23.46 [22.68 - 27.67] | 14.48 [13.99 - 17.15] |
| Consumables Increased to 75th Percentile | $6.91 [$6.04 - $8.16] | $27.03 [$23.21 - $34.70] | $42.71 [$36.67 - $54.83] | 254.61 [177.72 - 343.73] | 160.78 [112.11 - 217.18] |
| Consumables Increased to HIV levels | $8.26 [$8.19 - $10.75] | $77.77 [$69.50 - $79.03] | $122.88 [$109.80 - $124.87] | 109.53 [103.15 - 138.55] | 68.96 [64.91 - 87.32] |
| Consumables Increased to EPI Levels | $11.49 [$10.01 - $12.34] | $93.18 [$86.80 - $102.08] | $147.22 [$137.14 - $161.28] | 113.71 [106.57 - 131.24] | 71.6 [67.09 - 82.7] |
| **HSS Expansion Package** | **$15.59 [$14.44 - $17.63]** | **$2,233.31 [$2,222.41 - $2,246.85]** | **$3,528.64 [$3,511.42 - $3,550.02]** | **5.97 [5.48 - 6.85]** | **3.41 [3.1 - 3.97]** |
| HIV Program Scale-up Without HSS Expansion | $0.00 [$0.00 - $0.32] | $164.45 [$159.37 - $169.19] | $259.83 [$251.81 - $267.32] | -1.0 [-1.0 - 0.87] | -1.0 [-1.0 - 0.18] |
| TB Program Scale-up Without HSS Expansion | $1.45 [$0.07 - $2.80] | $3.07 [$-2.19 - $3.93] | $4.84 [$-3.47 - $6.21] | 541.3 [19.45 - 1120.74] | 342.96 [11.94 - 708.96] |
| Malaria Program Scale-up Without HSS Expansion | $3.91 [$1.17 - $4.67] | $547.03 [$543.43 - $554.21] | $864.31 [$858.61 - $875.66] | 6.14 [1.12 - 7.57] | 3.52 [0.34 - 4.43] |
| **HTM Programs Scale-up Without HSS Expansion** | **$5.38 [$3.70 - $6.14]** | **$720.19 [$717.70 - $727.18]** | **$1,137.90 [$1,133.96 - $1,148.95]** | **6.4 [4.13 - 7.55]** | **3.68 [2.25 - 4.41]** |
| HIV Program Scale-up With HSS Expansion Package | $18.18 [$17.15 - $20.09] | $1,984.39 [$1,972.54 - $1,993.00] | $3,135.34 [$3,116.61 - $3,148.95] | 8.12 [7.64 - 9.18] | 4.77 [4.47 - 5.45] |
| TB Program Scale-up With HSS Expansion Package | $17.59 [$17.19 - $19.34] | $2,235.77 [$2,224.87 - $2,249.78] | $3,532.52 [$3,515.30 - $3,554.65] | 6.87 [6.69 - 7.6] | 3.98 [3.87 - 4.44] |
| Malaria Program Scale-up With HSS Expansion Package | $16.34 [$15.12 - $17.70] | $2,759.94 [$2,752.59 - $2,774.92] | $4,360.71 [$4,349.10 - $4,384.37] | 4.92 [4.48 - 5.42] | 2.75 [2.47 - 3.07] |
| **HTM Programs Scale-up With HSS Expansion Package** | **$22.69 [$20.43 - $23.36]** | **$2,507.07 [$2,493.32 - $2,518.76]** | **$3,961.17 [$3,939.44 - $3,979.64]** | **8.1 [7.15 - 8.28]** | **4.76 [4.16 - 4.87]** |

**Supplementary Table B.2: Return on Investment by Scenario (Value of Statistical Life Year = $834; Health and cost discount rate = 5%)**

| **Scenario** | **Monetised health benefits ($, billion)** | **Service-level costs ($, million)** | **Above Service-level costs ($, million)**  *(assumed to be equal to 58% of service-level costs)* | **ROI (assuming zero above service level costs)** | **ROI (assuming non-zero above service level costs)** |
| --- | --- | --- | --- | --- | --- |
| Pessimistic HRH Scale-up | $8.94 [$7.33 - $9.78] | $780.68 [$774.59 - $788.20] | $1,233.47 [$1,223.86 - $1,245.35] | 10.55 [8.34 - 11.58] | 6.31 [4.91 - 6.96] |
| Historical HRH Scale-up | $8.49 [$6.39 - $8.61] | $678.02 [$672.73 - $679.46] | $1,071.28 [$1,062.92 - $1,073.54] | 11.49 [8.41 - 11.79] | 6.91 [4.96 - 7.1] |
| Optimistic HRH Scale-up | $12.36 [$10.84 - $13.47] | $1,368.15 [$1,366.44 - $1,377.23] | $2,161.67 [$2,158.97 - $2,176.02] | 8.04 [6.88 - 8.79] | 4.72 [3.99 - 5.19] |
| Primary Healthcare Workforce Scale-up | $5.92 [$5.36 - $6.66] | $272.85 [$270.28 - $286.09] | $431.10 [$427.05 - $452.02] | 19.62 [18.5 - 23.49] | 12.05 [11.34 - 14.5] |
| Consumables Increased to 75th Percentile | $5.18 [$4.75 - $6.56] | $24.46 [$21.14 - $31.58] | $38.64 [$33.41 - $49.89] | 210.94 [152.35 - 301.62] | 133.14 [96.06 - 190.53] |
| Consumables Increased to HIV levels | $6.52 [$6.25 - $8.73] | $71.31 [$63.83 - $72.61] | $112.68 [$100.85 - $114.73] | 91.13 [86.48 - 122.67] | 57.31 [54.37 - 77.27] |
| Consumables Increased to EPI Levels | $9.02 [$8.00 - $9.62] | $85.43 [$79.48 - $93.58] | $134.98 [$125.58 - $147.86] | 97.54 [93.78 - 114.24] | 61.37 [58.99 - 71.93] |
| **HSS Expansion Package** | **$11.56 [$10.94 - $13.30]** | **$2,028.46 [$2,018.40 - $2,040.54]** | **$3,204.97 [$3,189.08 - $3,224.06]** | **4.7 [4.4 - 5.52]** | **2.61 [2.42 - 3.13]** |
| HIV Program Scale-up Without HSS Expansion | $0.00 [$0.00 - $0.20] | $152.83 [$148.29 - $156.98] | $241.47 [$234.31 - $248.04] | -1.0 [-1.0 - 0.24] | -1.0 [-1.0 - -0.21] |
| TB Program Scale-up Without HSS Expansion | $1.36 [$0.03 - $2.45] | $2.81 [$-1.82 - $3.51] | $4.44 [$-2.88 - $5.55] | 604.68 [8.38 - 1209.13] | 382.34 [4.94 - 764.98] |
| Malaria Program Scale-up Without HSS Expansion | $3.00 [$0.81 - $3.46] | $497.65 [$494.40 - $503.86] | $786.29 [$781.16 - $796.10] | 5.04 [0.62 - 5.98] | 2.82 [0.02 - 3.42] |
| **HTM Programs Scale-up Without HSS Expansion** | **$4.19 [$2.92 - $4.79]** | **$658.22 [$655.85 - $664.26]** | **$1,039.98 [$1,036.25 - $1,049.54]** | **5.3 [3.44 - 6.29]** | **2.99 [1.81 - 3.61]** |
| HIV Program Scale-up With HSS Expansion Package | $13.51 [$12.60 - $15.21] | $1,802.16 [$1,791.52 - $1,809.97] | $2,847.42 [$2,830.60 - $2,859.75] | 6.48 [5.99 - 7.49] | 3.74 [3.42 - 4.37] |
| TB Program Scale-up With HSS Expansion Package | $13.49 [$12.75 - $14.60] | $2,030.63 [$2,020.84 - $2,043.22] | $3,208.40 [$3,192.93 - $3,228.28] | 5.64 [5.28 - 6.15] | 3.21 [2.97 - 3.53] |
| Malaria Program Scale-up With HSS Expansion Package | $12.41 [$11.17 - $13.39] | $2,507.90 [$2,501.07 - $2,521.19] | $3,962.48 [$3,951.69 - $3,983.48] | 3.92 [3.45 - 4.35] | 2.11 [1.82 - 2.39] |
| **HTM Programs Scale-up With HSS Expansion Package** | **$17.16 [$15.26 - $17.83]** | **$2,278.13 [$2,265.65 - $2,288.68]** | **$3,599.45 [$3,579.72 - $3,616.11]** | **6.58 [5.7 - 6.79]** | **3.8 [3.24 - 3.93]** |

2. **Alternative VSLY**

Supplementary Figure B4 and Supplementary Tables B3 and B4 present ROI estimates for the joint HTM scenario under the three approaches assuming alternative VSLY of $425.96 and $2427.31, applying a 3% discount rate for both health and cost. With the lower VSLY assumption, the threshold incremental above service level costs for the diagonal approach to provide a greater ROI than the vertical approach under the two above service level cost assumptions for the vertical approach (equal to 0% of service level cost and 58% of service level cost) were $508.72 (20.29% of its own incremental service level cost or 2.85% of the total project health spending for 2025-2035) and $1115.99 million (44.51% of its own incremental service level cost or 6.25% of the total project health spending for 2025-2035), respectively. With the higher VSLY assumption, the threshold incremental above service level costs for the diagonal approach to provide a greater ROI than the vertical approach under the two above service level cost assumptions were $508.72 (20.29% of its own incremental service level cost or 2.85% of the total project health spending for 2025-2035) and $1694.88 million (67.6% of its own incremental service level cost or 9.5% of the total project health spending for 2025-2035).

**Supplementary Figure B.4 Return on Investment (ROI) assuming alternative estimates of Value of Statistical Life Year (VSLY)**

| 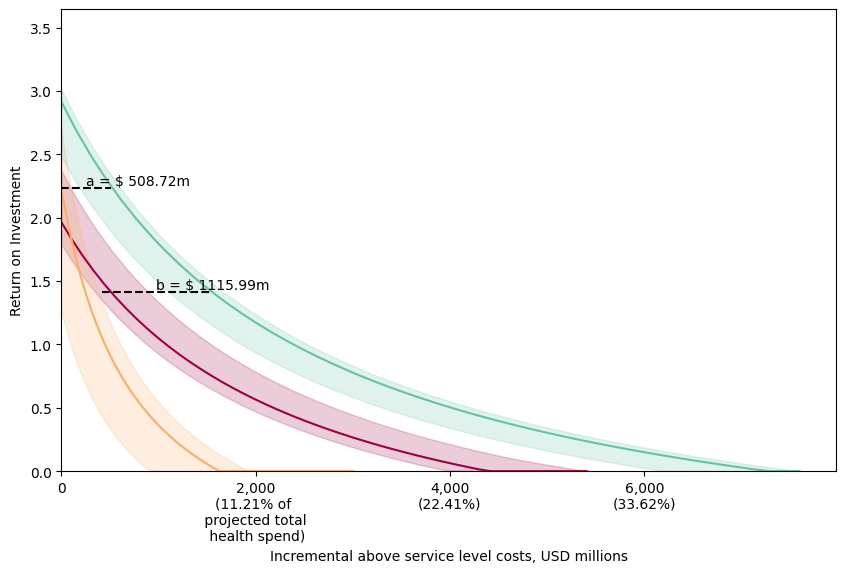 |
| --- |
| (i) VSLY = $425.96  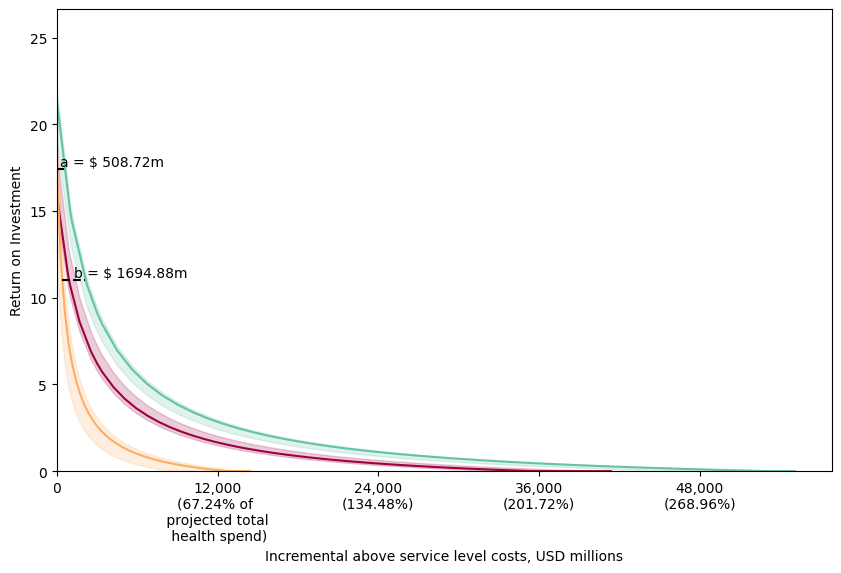 |
| (ii) VSLY = $2,427.31  * Note: ROI estimates are clipped at a minimum value of 0. A 3% discount rate is applied to both health  and costs. |

**Supplementary Table B.3: Return on Investment by Scenario (Value of Statistical Life Year = $425.96; Health and cost discount rate = 3%)**

| **Scenario** | **Monetised health benefits ($, billion)** | **Service-level costs ($, million)** | **Above Service-level costs ($, million)**  *(assumed to be equal to 58% of service-level costs)* | **ROI (assuming zero above service level costs)** | **ROI (assuming non-zero above service level costs)** |
| --- | --- | --- | --- | --- | --- |
| Pessimistic HRH Scale-up | $5.02 [$4.20 - $5.53] | $854.19 [$847.36 - $862.40] | $1,349.63 [$1,338.83 - $1,362.59] | 4.92 [3.89 - 5.5] | 2.75 [2.09 - 3.11] |
| Historical HRH Scale-up | $4.77 [$3.62 - $4.83] | $739.51 [$733.69 - $741.12] | $1,168.43 [$1,159.22 - $1,170.97] | 5.48 [3.88 - 5.58] | 3.1 [2.09 - 3.16] |
| Optimistic HRH Scale-up | $6.98 [$6.22 - $7.67] | $1,510.32 [$1,508.64 - $1,520.49] | $2,386.31 [$2,383.66 - $2,402.37] | 3.63 [3.1 - 4.05] | 1.93 [1.59 - 2.2] |
| Primary Healthcare Workforce Scale-up | $3.35 [$3.05 - $3.69] | $297.86 [$295.07 - $312.54] | $470.62 [$466.20 - $493.82] | 9.66 [9.16 - 11.51] | 5.75 [5.43 - 6.92] |
| Consumables Increased to 75th Percentile | $2.96 [$2.66 - $3.64] | $27.03 [$23.21 - $34.70] | $42.71 [$36.67 - $54.83] | 108.5 [77.3 - 152.71] | 68.3 [48.56 - 96.29] |
| Consumables Increased to HIV levels | $3.64 [$3.54 - $4.83] | $77.77 [$69.50 - $79.03] | $122.88 [$109.80 - $124.87] | 46.79 [44.14 - 61.66] | 29.24 [27.57 - 38.66] |
| Consumables Increased to EPI Levels | $5.05 [$4.49 - $5.40] | $93.18 [$86.80 - $102.08] | $147.22 [$137.14 - $161.28] | 49.45 [47.49 - 57.73] | 30.93 [29.69 - 36.17] |
| **HSS Expansion Package** | **$6.62 [$6.22 - $7.57]** | **$2,233.31 [$2,222.41 - $2,246.85]** | **$3,528.64 [$3,511.42 - $3,550.02]** | **1.97 [1.79 - 2.37]** | **0.88 [0.77 - 1.13]** |
| HIV Program Scale-up Without HSS Expansion | $0.00 [$0.00 - $0.12] | $164.45 [$159.37 - $169.19] | $259.83 [$251.81 - $267.32] | -1.0 [-1.0 - -0.28] | -1.0 [-1.0 - -0.55] |
| TB Program Scale-up Without HSS Expansion | $0.72 [$0.02 - $1.29] | $3.07 [$-2.19 - $3.93] | $4.84 [$-3.47 - $6.21] | 267.02 [5.49 - 546.64] | 169.37 [3.11 - 345.61] |
| Malaria Program Scale-up Without HSS Expansion | $1.70 [$0.48 - $1.98] | $547.03 [$543.43 - $554.21] | $864.31 [$858.61 - $875.66] | 2.1 [-0.13 - 2.64] | 0.96 [-0.45 - 1.3] |
| **HTM Programs Scale-up Without HSS Expansion** | **$2.35 [$1.63 - $2.66]** | **$720.19 [$717.70 - $727.18]** | **$1,137.90 [$1,133.96 - $1,148.95]** | **2.24 [1.27 - 2.7]** | **1.05 [0.43 - 1.34]** |
| HIV Program Scale-up With HSS Expansion Package | $7.74 [$7.25 - $8.65] | $1,984.39 [$1,972.54 - $1,993.00] | $3,135.34 [$3,116.61 - $3,148.95] | 2.89 [2.65 - 3.38] | 1.46 [1.31 - 1.77] |
| TB Program Scale-up With HSS Expansion Package | $7.63 [$7.31 - $8.31] | $2,235.77 [$2,224.87 - $2,249.78] | $3,532.52 [$3,515.30 - $3,554.65] | 2.41 [2.27 - 2.7] | 1.16 [1.07 - 1.34] |
| Malaria Program Scale-up With HSS Expansion Package | $7.07 [$6.41 - $7.61] | $2,759.94 [$2,752.59 - $2,774.92] | $4,360.71 [$4,349.10 - $4,384.37] | 1.55 [1.32 - 1.76] | 0.61 [0.47 - 0.75] |
| **HTM Programs Scale-up With HSS Expansion Package** | **$9.76 [$8.72 - $10.10]** | **$2,507.07 [$2,493.32 - $2,518.76]** | **$3,961.17 [$3,939.44 - $3,979.64]** | **2.92 [2.48 - 3.01]** | **1.48 [1.2 - 1.54]** |

**Table B.4: Return on Investment by Scenario (Value of Statistical Life Year = $2,427.31; Health and cost discount rate = 3%)**

| **Scenario** | **Monetised health benefits ($, billion)** | **Service-level costs ($, million)** | **Above Service-level costs ($, million)**  *(assumed to be equal to 58% of service-level costs)* | **ROI (assuming zero above service level costs)** | **ROI (assuming non-zero above service level costs)** |
| --- | --- | --- | --- | --- | --- |
| Pessimistic HRH Scale-up | $28.58 [$23.93 - $31.50] | $854.19 [$847.36 - $862.40] | $1,349.63 [$1,338.83 - $1,362.59] | 32.74 [26.86 - 36.04] | 20.35 [16.63 - 22.44] |
| Historical HRH Scale-up | $27.17 [$20.60 - $27.54] | $739.51 [$733.69 - $741.12] | $1,168.43 [$1,159.22 - $1,170.97] | 35.91 [26.83 - 36.48] | 22.36 [16.62 - 22.72] |
| Optimistic HRH Scale-up | $39.80 [$35.46 - $43.71] | $1,510.32 [$1,508.64 - $1,520.49] | $2,386.31 [$2,383.66 - $2,402.37] | 25.38 [22.36 - 27.77] | 15.7 [13.79 - 17.21] |
| Primary Healthcare Workforce Scale-up | $19.06 [$17.38 - $21.04] | $297.86 [$295.07 - $312.54] | $470.62 [$466.20 - $493.82] | 59.73 [56.92 - 70.29] | 37.44 [35.66 - 44.12] |
| Consumables Increased to 75th Percentile | $16.87 [$15.15 - $20.74] | $27.03 [$23.21 - $34.70] | $42.71 [$36.67 - $54.83] | 622.96 [445.2 - 874.91] | 393.91 [281.4 - 553.38] |
| Consumables Increased to HIV levels | $20.76 [$20.20 - $27.50] | $77.77 [$69.50 - $79.03] | $122.88 [$109.80 - $124.87] | 271.31 [256.21 - 356.09] | 171.35 [161.79 - 225.01] |
| Consumables Increased to EPI Levels | $28.79 [$25.57 - $30.79] | $93.18 [$86.80 - $102.08] | $147.22 [$137.14 - $161.28] | 286.5 [275.32 - 333.66] | 180.96 [173.88 - 210.81] |
| **HSS Expansion Package** | **$37.74 [$35.42 - $43.16]** | **$2,233.31 [$2,222.41 - $2,246.85]** | **$3,528.64 [$3,511.42 - $3,550.02]** | **15.9 [14.9 - 18.22]** | **9.7 [9.06 - 11.16]** |
| HIV Program Scale-up Without HSS Expansion | $0.00 [$0.00 - $0.69] | $164.45 [$159.37 - $169.19] | $259.83 [$251.81 - $267.32] | -1.0 [-1.0 - 3.08] | -1.0 [-1.0 - 1.58] |
| TB Program Scale-up Without HSS Expansion | $4.08 [$0.13 - $7.35] | $3.07 [$-2.19 - $3.93] | $4.84 [$-3.47 - $6.21] | 1516.91 [36.01 - 3119.68] | 960.44 [22.42 - 1974.12] |
| Malaria Program Scale-up Without HSS Expansion | $9.66 [$2.72 - $11.30] | $547.03 [$543.43 - $554.21] | $864.31 [$858.61 - $875.66] | 16.67 [3.93 - 19.73] | 10.18 [2.12 - 12.12] |
| **HTM Programs Scale-up Without HSS Expansion** | **$13.41 [$9.31 - $15.14]** | **$720.19 [$717.70 - $727.18]** | **$1,137.90 [$1,133.96 - $1,148.95]** | **17.44 [11.91 - 20.06]** | **10.67 [7.17 - 12.33]** |
| HIV Program Scale-up With HSS Expansion Package | $44.11 [$41.33 - $49.27] | $1,984.39 [$1,972.54 - $1,993.00] | $3,135.34 [$3,116.61 - $3,148.95] | 21.16 [19.82 - 23.98] | 13.02 [12.18 - 14.81] |
| TB Program Scale-up With HSS Expansion Package | $43.47 [$41.63 - $47.36] | $2,235.77 [$2,224.87 - $2,249.78] | $3,532.52 [$3,515.30 - $3,554.65] | 18.45 [17.62 - 20.06] | 11.31 [10.78 - 12.33] |
| Malaria Program Scale-up With HSS Expansion Package | $40.28 [$36.54 - $43.37] | $2,759.94 [$2,752.59 - $2,774.92] | $4,360.71 [$4,349.10 - $4,384.37] | 13.51 [12.24 - 14.75] | 8.18 [7.38 - 8.97] |
| **HTM Programs Scale-up With HSS Expansion Package** | **$55.62 [$49.70 - $57.58]** | **$2,507.07 [$2,493.32 - $2,518.76]** | **$3,961.17 [$3,939.44 - $3,979.64]** | **21.32 [18.83 - 21.86]** | **13.12 [11.55 - 13.47]** |

4.	National Planning Commission. *Medium and Long-Term Impacts of a Moderate Lockdown (Social Restrictions) in Response to the COVID-19 Pandemic in Malawi: A Rapid Cost-Benefit Analysis*. https://afidep.org/publication/medium-and-long-term-impacts-of-a-moderate-lockdown-social-restrictions-in-response-to-the-covid-19-pandemic-in-malawi-a-rapid-cost-benefit-analysis/.

5.	Robinson, L. A. *et al.* Reference Case Guidelines for Benefit-Cost Analysis in Global Health and Development. SSRN Scholarly Paper at https://doi.org/10.2139/ssrn.4015886 (2019).

6.	Lomas, J., Claxton, K. & Ochalek, J. Accounting for country- and time-specific values in the economic evaluation of health-related projects relevant to low- and middle-income countries. *Health Policy Plan.* **37**, 45–54 (2022).

7.	Eayres, D. & Williams, E. S. Evaluation of methodologies for small area life expectancy estimation. *J. Epidemiol. Community Health* **58**, 243–249 (2004).

8.	The World Bank. World Development Indicators. https://databank.worldbank.org/source/world-development-indicators.

9.	Haacker, M., Hallett, T. B. & Atun, R. On discount rates for economic evaluations in global health. *Health Policy Plan.* **35**, 107–114 (2020).

10.	Bertram, M. Y., Lauer, J. A., Stenberg, K. & Edejer, T. T. T. Methods for the Economic Evaluation of Health Care Interventions for Priority Setting in the Health System: An Update From WHO CHOICE. *Int. J. Health Policy Manag.* **10**, 673–677 (2021).
